# Neutrophil-primed immunopathology in poorly-controlled diabetes worsens matrix destruction in pulmonary tuberculosis

**DOI:** 10.64898/2026.05.24.26353970

**Authors:** Pei Min Thong, Ting Huey Hu, Justin S.G. Ooi, Fei Kean Loh, Hyeyoung Lee, Chen Bai, Hai Tarng Chong, Anabel Jia Wei Chang, Caroline Victoria Choong, Lovel Galamay, Darius L.L. Beh, Alicia X.Y. Ang, Lionel H.W. Lum, Samantha Peiling Yang, Amanda Yuan Ling Lim, Shao Feng Mok, Andres F Vallejo, Shih Ling Kao, Kuan Rong Chan, Catherine W.M. Ong

## Abstract

**Background:** Diabetes mellitus (DM) worsens pulmonary tuberculosis (TB) and drives systemic hyper-inflammation, but the underlying mechanisms remain unknown. Neutrophils have key roles in TB immunopathology and lung cavitation. Here, we determine the role of neutrophils in DMTB patients and in driving TB immunopathology.

**Methods:** Sputum and plasma from 30 TB and 30 DMTB patients were analysed for proteases and cytokines using Luminex bead array. Whole blood transcriptomics identified transcriptional differences. Single-cell RNA sequencing characterised neutrophil subsets and dysregulated pathways. Neutrophil function of poorly-controlled DM patients (HbA1c>8%) and healthy controls (HC) were examined following *Mycobacterium tuberculosis* stimulation, including reactive oxygen species (ROS), neutrophil extracellular traps (NETs), and phagocytosis. Pathways were interrogated using chemical inhibitors, protein array and western blot.

**Results:** Compared to non-diabetic TB patients, poorly-controlled DMTB patients showed up-regulated sputum MMP-8 and MMP-9, associated with increased collagen-destruction and lung cavity formation. Circulating neutrophil count and neutrophil-derived plasma MMP-8 were up-regulated, alongside transcriptional enrichment of extracellular matrix degradation and inflammatory pathways including TNF and RAGE. Single-cell profiling identified reduced cycling neutrophil subset and myelocytes in DMTB, with overall reduced antibacterial and cell-killing signatures. *Ex vivo* mycobacterial stimulation of DM neutrophils increased ROS and MMP-9 with impaired NETs and delayed phagocytosis. TNFR1, TNFR2, and RAGE were up-regulated. RAGE inhibition with rosiglitazone mitigated *Mtb*-induced ROS and MMP-8 release.

**Conclusion:** DM worsens neutrophil-driven tissue destruction and inflammation in TB via dysregulated TNF and RAGE-signalling, priming neutrophils towards immunopathology. Targeting RAGE alongside tight glycaemic control may dampen neutrophil hyper-inflammatory responses to limit tissue destruction.

## Introduction

The global prevalence of diabetes mellitus (DM) among pulmonary tuberculosis (TB) patients is rising (1). A systematic review and meta-analysis of studies conducted between 1980 and 2020 reported a DM prevalence of 21% among TB patients in South Asia, a region that accounts for nearly 44% of global TB cases, highlighting the accelerating burden of this twin epidemic (2). DM patients have a two-fold higher risk of developing TB infection and TB disease, and an increase in DM and TB co-infections is expected (3). Concurrent DM and TB result in severe clinical manifestations, including more lung cavitations (4), higher mycobacterial burden (5) and longer periods of infectiousness. Delayed sputum culture sterilisation in these patients could heighten the risk of community TB transmission due to slower mycobacterial clearance (6), posing a potential setback to global TB control efforts. Therefore, understanding the interaction between DM and TB is essential to mitigate this rising burden and to develop targeted interventions for improved outcomes.

Immunopathology of DMTB is characterised by systemic hyperinflammation (5, 7, 8) and extensive pulmonary destruction (9–11). Whole blood transcriptomic studies identified an increase in neutrophil and innate immune pathway activation in DMTB (12), and delayed resolution of inflammation (7). This finding is supported by a longitudinal study that monitored systemic cytokines in DMTB patients during anti-TB treatment, showing persistent inflammation in these patients (5). This persistent pro-inflammatory state in DM patients is likely due to ongoing bacterial replication or impaired bacterial clearance, implied by correlations between whole blood transcriptomic data and lung inflammation (13). However, the mechanisms linking this systemic immune dysregulation to pulmonary tissue damage remain unclear.

Excessive neutrophil accumulation in the lungs is detrimental to *Mycobacterium tuberculosis (Mtb)* infection (14–16) and associated with increased TB-attributed mortality (17). Neutrophils are a major source of proteolytic enzymes, including matrix metalloproteinases (MMPs) such as MMP-8 and MMP-9, which drive extracellular matrix degradation and tissue remodelling (18). An imbalance between MMPs and their inhibitors, tissue inhibitors of metalloproteinases (TIMPs), is associated with a matrix-degrading phenotype in pulmonary TB (18). Although upregulated systemic MMPs were reported in DMTB patients compared to TB alone (19), their role and drivers within the respiratory compartment remains unclear. Building on transcriptomic evidence identifying increased neutrophil pathway activation in DMTB, we determine neutrophil function alongside protease activity and cytokine expression at both the protein and gene levels in a DMTB model and patient samples to elucidate mechanisms underlying immune dysregulation in DMTB.

## Methods

### Study Design and Participants

This study was conducted at the National University of Singapore, National University Hospital, and National Tuberculosis Care Centre. Ethics approval was obtained from the NUS Institutional Review Board (Ref: LH-19-001) and NHG Domain Specific Review Board (Ref: 2018/01213). A total of 134 participants were recruited between 2018 and 2023; these comprised 35 healthy controls (HC), 34 poorly-controlled DM patients with HbA1c >8%, 35 non-diabetic TB patients and 30 DMTB patients (10 with HbA1c 6.5–8%, and 20 poorly-controlled DM of HbA1c>8%). The diagnosis of pulmonary TB was microbiologically confirmed by TB culture and/or GeneXpert, and all patients with PTB were within 7 days of TB treatment initiation. Exclusion criteria included Human immunodeficiency virus coinfection, prior TB disease, pre-existing lung disease, and immunosuppressive therapy. Detailed descriptions of the study design and experimental procedures are provided in the Supplementary Materials.

### Chest radiograph scoring

Chest radiographs were scored for cavitation and disease extent by three blinded clinicians (CWMO, AXYA and DLLB) using a validated scoring system (20).

### Sample Collection and Processing

Whole blood was collected in Tempus tubes for RNA extraction and EDTA tubes for neutrophil isolation and plasma separation. Primary neutrophils were isolated using EasySep™ Direct Human Neutrophil Isolation Kit (Stemcell Technologies, #19666). Sputum was collected and processed in BSL-3 facility as previously described (21). Respiratory samples and plasma were sterile-filtered as per local BSL3 regulations prior analysis (22)

### Plasma and Sputum Protein Analysis

Cytokines, matrix metalloproteinases (MMPs), and neutrophil markers in plasma and sputum were measured using Luminex assays and ELISA kits (R&D Systems). Sputum protein levels were normalised to total protein concentration. Type I collagen and elastin degradation were assessed using EnzChek® Collagenase and Elastase assay kits (Invitrogen) (23).

Sputum MMP-9 activity was assessed by gelatin zymography on 11% SDS-PAGE gels with 1.2% gelatin (24). Gels were incubated in collagenase buffer at 37°C for 40 hours. Recombinant MMP-9 (500pg) served as a positive control. Band intensity was quantified using FIJI (ImageJ).

### Whole blood bulk RNA sequencing

RNA was extracted from whole blood using TRIzol and column purification. Libraries were prepared with poly(A) capture and sequenced on an Illumina NovaSeq 6000 (150 bp paired-end). Differential gene expression was analysed using edgeR (25) and Limma (26). Gene enrichment analysis was performed with ShinyGO (27). Immune cell deconvolution was performed using CIBERSORTx (28, 29), and gene co-expression networks were constructed using Weighted Gene Co-expression Network Analysis (WGCNA) to identify functional gene modules (30, 31).

### Single-cell RNA data processing and analysis

Freshly isolated neutrophils were processed using the 10x Genomics Chromium Fixed RNA Profiling Kit and libraries were sequenced on a DNBSEQ T7 platform (BGI). Raw sequencing data were processed using the Cell Ranger multi pipeline (v9.0.1, 10x Genomics). The filtered gene-barcode matrix of UMI counts was analysed with Seurat V5 (32) for quality control, normalization, dimensionality reduction, integration, clustering, and visualization. Principal component analysis (PCA) followed by uniform manifold approximation and projection (UMAP) was used for dimensionality reduction. Cell clustering was performed using the shared nearest neighbor (SNN) graph-based Louvain algorithm. Cell clusters were annotated using established neutrophil marker genes (33) and non-neutrophil cell clusters were excluded from further analysis. Differential gene expression between DMTB and TB samples was identified using the FindMarkers function. Functional enrichment analysis was performed using the enrichGO function from the clusterProfiler package. Data visualization was performed using ggplot2, Seurat, and scCustomize. Detailed quality control thresholds and analysis parameters are provided in the Supplementary Methods.

### Neutrophil Functional Assays

Primary neutrophils from DM patients and healthy controls were concurrently stimulated as a set with *Mycobacterium tuberculosis (Mtb)* H37Rv at multiplicity of infectionLofL10 (22). Intracellular and extracellular reactive oxygen species (ROS) were measured using luminol and isoluminol-based chemiluminescence (34). Phagocytosis of mCherry-labelled BCG was assessed by flow cytometry, with the gating strategy shown in Supplementary Method. NETs were quantified by dsDNA release and neutrophil elastase-DNA ELISA.

### Neutrophil Cell Signalling Analysis

NF-κB pathway activation was assessed using Human NF-κB signalling arrays (R&D systems, #ARY029) following stimulation with heat-killed *Mtb*. Western blotting was performed on lysates from live *Mtb*-stimulated neutrophils to evaluate receptor for advanced glycation end product (RAGE) (Invitrogen, #PA5-24787), tumour necrosis factor receptors TNFR1 (Cell Signalling Technology, #3736S), and TNFR2 (Cell Signalling Technology, #72337) protein expression.

### Statistical Analysis

All statistical analyses were performed using GraphPad Prism v10. Data are presented as median with interquartile range unless otherwise stated. Data were assessed for normality prior to analysis. For normally distributed data, comparisons between two groups were performed using an unpaired t-test, while comparisons among multiple groups were conducted using one-way ANOVA or two-way ANOVA followed by Sidak’s multiple comparisons test. For non-normally distributed data, the Mann–Whitney test was used for two-group comparisons, and the Kruskal-Wallis test with Dunn’s multiple comparisons test was applied for multiple-group analyses. Paired tests were used for paired HC-DM *ex vivo* comparisons. A *p*-value <0.05 was considered statistically significant.

## Results

### Baseline characteristics of study participants

The baseline characteristics of the 134 participants are in **Table□1**. There were 79 males (59%), with higher predominance in the TB (71.4%) and DMTB (76.7%) groups. Within DMTB, 20 had poorly-controlled DM (HbA1c>8%), and 10 had controlled DM (HbA1c 6.5-8%). Median acid-fast bacilli (AFB) smear scores showed a numerical increase from TB patients without DM to those with controlled DM (C_DMTB) and poorly-controlled DM (PC_DMTB).

**Table 1:** Study demographics.

| Characteristics | HC (n=35) | DM (n=34) | TB (n=35) | DMTB (n=30) |  | p-values |
| --- | --- | --- | --- | --- | --- | --- |
|  |  |  |  | C_DMTB (n=10) | PC_DMTB (n=20) |  |
| Median age, ^ years (IQR) | 35 (31-46) | 49 (37-55) | 39 (28-49) | 60 (51-72) | 46 (37-62) | <0.0001 |
| Males (%) | 15 (42.9%) | 16 (47.1%) | 25 (71.4%) | 6 (60%) | 17 (85%) | 0.0087 |
| Median HbA1c, % (IQR)* | 5.3 (5.1-5.4) | 9.5 (8.8-10.9) | 5.6 (5.1-5.8) | 7.1 (6.8-7.6) | 12.1 (10.1-13.7) | <0.0001 |
| Median BMI, kg/m<sup>2</sup> (IQR)# | 22.3 (20.3-24.5) | 31.3 (26.2-34.6) | 20.9 (18.5-23.3) | 22.7 (17.9-27.0) | 22.0 (19.9-24.5) | <0.0001 |
| Median AFB smear (IQR) | N.A. | N.A. | 1.0 (0-3) | 2.0 (0.75-3) | 2.5 (0-3) | 0.5141 |

Data is presented as median and interquartile range.
p-values are determined by Kruskal Wallis test or Fisher's exact test.
HC: healthy controls, DM: diabetes mellitus, TB: tuberculosis, C\_DMTB: TB patients with controlled DM, PC\_DMTB: TB patients with poorly-controlled DM. N.A.: not applicable.
AFB: Acid fast bacilli
^ $p < 0.05$ for comparison between HC & C\_DMTB, HC & PC\_DMTB, and TB & C\_DMTB
\* HbA1c measurements were not available for 5 HC; these participants self-reported no history of diabetes mellitus. $p < 0.05$ for comparison between HC & DM, HC & C\_DMTB, HC & PC\_DMTB, DM & TB, TB & PC\_DMTB, and C\_DMTB & PC\_DMTB,

### Up-regulated respiratory MMP-8 and MMP-9 is significantly associated with increased lung tissue destruction in poorly-controlled DMTB patients

Respiratory MMP phenotyping revealed significantly elevated concentrations of MMP-8 (7.48-fold) and MMP-9 (2.38-fold) in poorly-controlled DMTB patients compared to non-diabetic TB patients (both *p*<0.01, **Figure 1A**), while concentrations in controlled DMTB patients remained comparable to non-diabetic TB patients. Sputum collagen destruction was also increased in poorly-controlled DMTB patients (1.8-fold higher, *p*<0.01, **Figure 1B**) and strongly correlated with MMP-8 concentration (r=0.7343, *p*<0.0001, **Figure 1C**), implicating MMP-8 as the primary driver of type I collagen destruction in the respiratory compartment. The other collagenases MMP-1 and MMP-13, showed a weak positive and a moderate negative association, respectively (*p*<0.05, **Table S1**). Sputum MMP-9 was similarly strongly associated with elastase destruction (r=0.7109, *p*<0.0001, **Figure 1D**), and gelatin zymography confirmed increased active MMP-9 in poorly-controlled DMTB (**Figure 1E**). Although MMP inhibitor TIMP-2 (endogenous inhibitor of MMP-2 and –9) was also upregulated in poorly-controlled DMTB, TIMP-1 (a broad inhibitor of soluble MMPs including MMP-1, –2, –3, –7, –8 and –9) remained unchanged (**Figure S1**), functionally there is net matrix-destruction as evidenced by both collagen and elastase activity (35).

**Figure 1.**
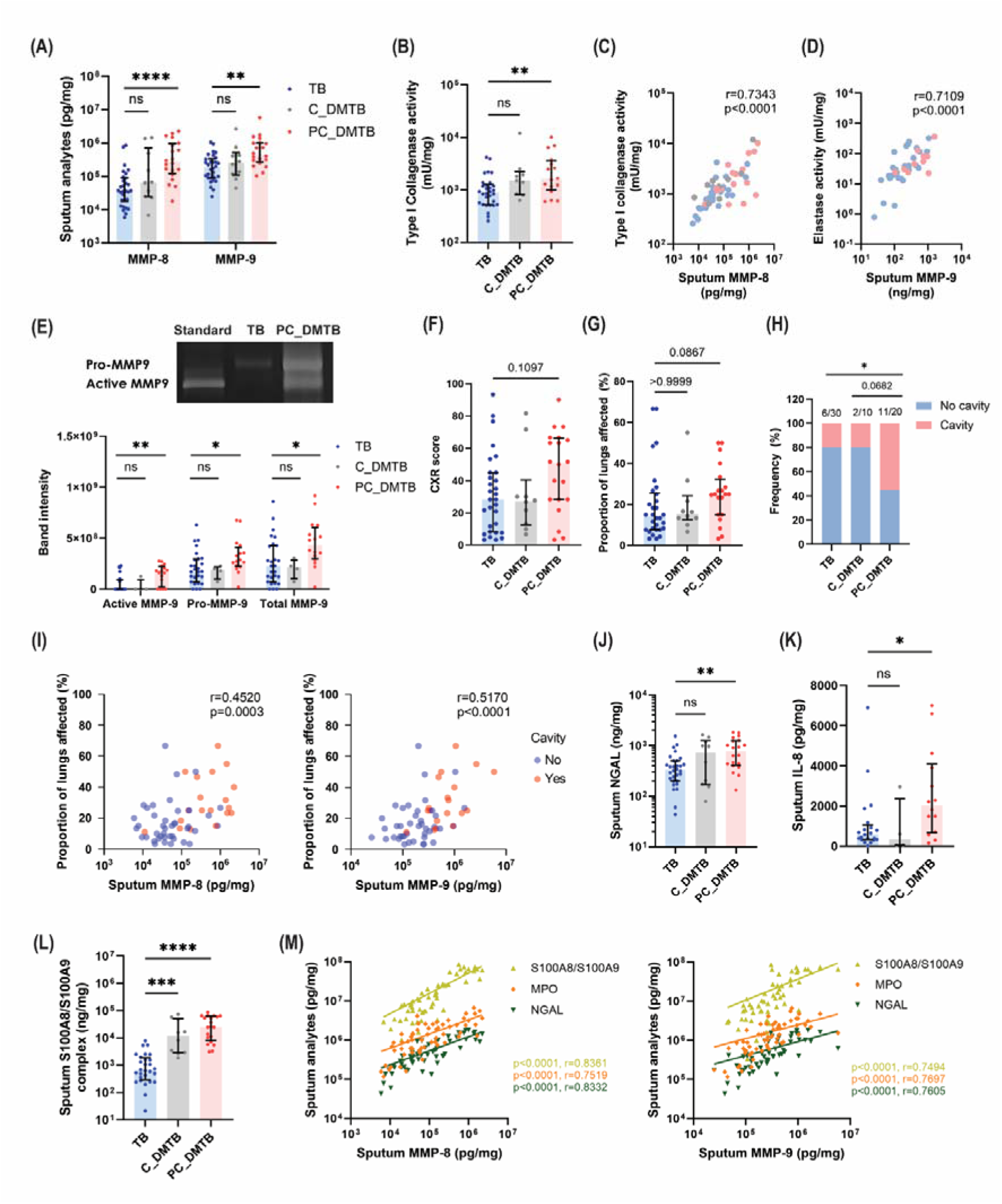
Increased neutrophil-derived respiratory MMP-8 and –9 concentrations and neutrophil factors correlate with worse lung pathology in pulmonary TB patients with poorly-controlled DM. Increased sputum (A) MMP-8 and MMP-9 concentrations and (B) type I collagenase activity in TB patients with poorly-controlled DM compared to TB patients without DM. Strong positive association between (C) sputum MMP-8 with type I collagenase activity and (D) sputum MMP-9 with elastase activity, indicate that MMP-8 and MMP-9 are key matrix metalloproteinases driving ECM degradation in lungs. Pink dots, PC_DMTB; grey dots, C_DMTB; blue dots, non-diabetic TB. (E) Gelatin zymogram show increased active MMP-9 in sputum in poorly-controlled DMTB patients. Higher (F) CXR score and (G) proportion of lungs affected in TB patients with poorly-controlled DM. (H) Higher frequency of cavity formation in TB patients with poorly-controlled DM. (I) Positive association between sputum MMP-8 and MMP-9 concentrations with extent of lung involvement in pulmonary TB patients. Patients with lung cavities had higher sputum MMPs in the sputum indicated by red dots clustering. (J) Neutrophil gelatinase-associated lipocalin (NGAL), (K) IL-8, and (L) S100A8/S100A9 proteins were increased in the sputum from TB patients with poorly-controlled DM relative to TB patients without DM. (M) Significant positive correlation between sputum MMP-8 and MMP-9 concentrations with neutrophils factors S100A8/S100A9 proteins, myeloperoxidase (MPO) and NGAL, indicating that these MMPs are neutrophil-derived. (A-B), (E), (F), (G), (J), (K) and (L) Kruskal Wallis test with Dunn’s multiple comparison test was performed. Data are presented as median and interquartile range. (C), (D), (I) and (M) Spearman correlation test was performed. (H) Fisher’s exact test was performed. n=30 TB, n=10 C_DMTB and n=20 PC_DMTB. \**p*<0.05, \*\**p*<0.01, \*\*\**p*<0.001, \*\*\*\**p*<0.0001

Although CXR showed no differences in total scores or lung involvement (**Figures 1F-G**), lung cavitation was significantly more frequent in poorly-controlled DMTB compared to non-diabetic TB patients (55% vs 20%, *p<*0.05, **Figure 1H**), while patients with controlled DMTB showed a similar frequency to non-diabetic patients with TB. Sputum MMP-8 and MMP-9 concentrations were associated with radiographic severity (both *p*<0.01, **Figure 1I**), linking MMP upregulation to tissue pathology. In parallel, neutrophil-associated inflammatory mediators (NGAL, IL-8 and S100A8/A9) were also significantly increased in the respiratory compartment of poorly-controlled DMTB patients and strongly associated with sputum MMP-8 and 9 concentrations (**Figure 1J-M**), supporting neutrophils as the primary source of proteolytic activity.

Next, we determined if use of anti-diabetic treatment affected MMP secretion. Several retrospective studies have reported potential benefits of metformin in DMTB, including improved outcomes, faster sputum culture conversion, and reduced relapse and cavitation (36–38). However, findings remain inconsistent, with some studies reporting no effect on culture conversion but reduced inflammatory responses (39). In our cohort, lung cavity formation was independent of metformin use, and neither metformin nor sulphonylurea modified sputum or plasma MMP-8 and MMP-9 concentrations (**Figure S2A-E**). *In vitro*, metformin did not alter neutrophil MMP-8 and MMP-9 release from *Mtb* stimulated-healthy neutrophils (**Figure S2F-G**), suggesting it does not directly modulate neutrophil-derived MMP secretion in this context.

Overall, these findings suggest that poor glycaemic control is associated with a more pronounced inflammatory and tissue-destructive phenotype, whereas patients with controlled DM with TB closely resembled non-diabetic TB patients, with no significant differences observed between the two groups.

### Whole blood transcriptomics implicate neutrophils in DMTB pathogenesis

Given the marked increase in respiratory MMPs and neutrophil factors in poorly-controlled DMTB patients, we determined if similar neutrophil signatures are reflected systemically. Principal Component Analysis (PCA) showed clear separation between HC and patient groups (**Figure 2A**). Differential expression analysis identified 462 genes significantly upregulated in poorly-controlled DMTB compared with TB patients, including ECM-associated gene *COL4A2* (**Figure 2B**). Reactome analysis revealed significant enrichment of collagen degradation and ECM organisation pathways (**Figure 2C**), consistent with respiratory MMP findings of increased ECM destruction which was more marked in poorly-controlled diabetic patients. WGCNA analysis identified a module (black) strongly correlated with poorly-controlled DMTB (r=0.77, *p*<0.05, **Figure 2D**), which increased with AFB burden (**Figure 2E**), and was enriched for ECM-related pathways (**Figure 2F**).

**Figure 2.**
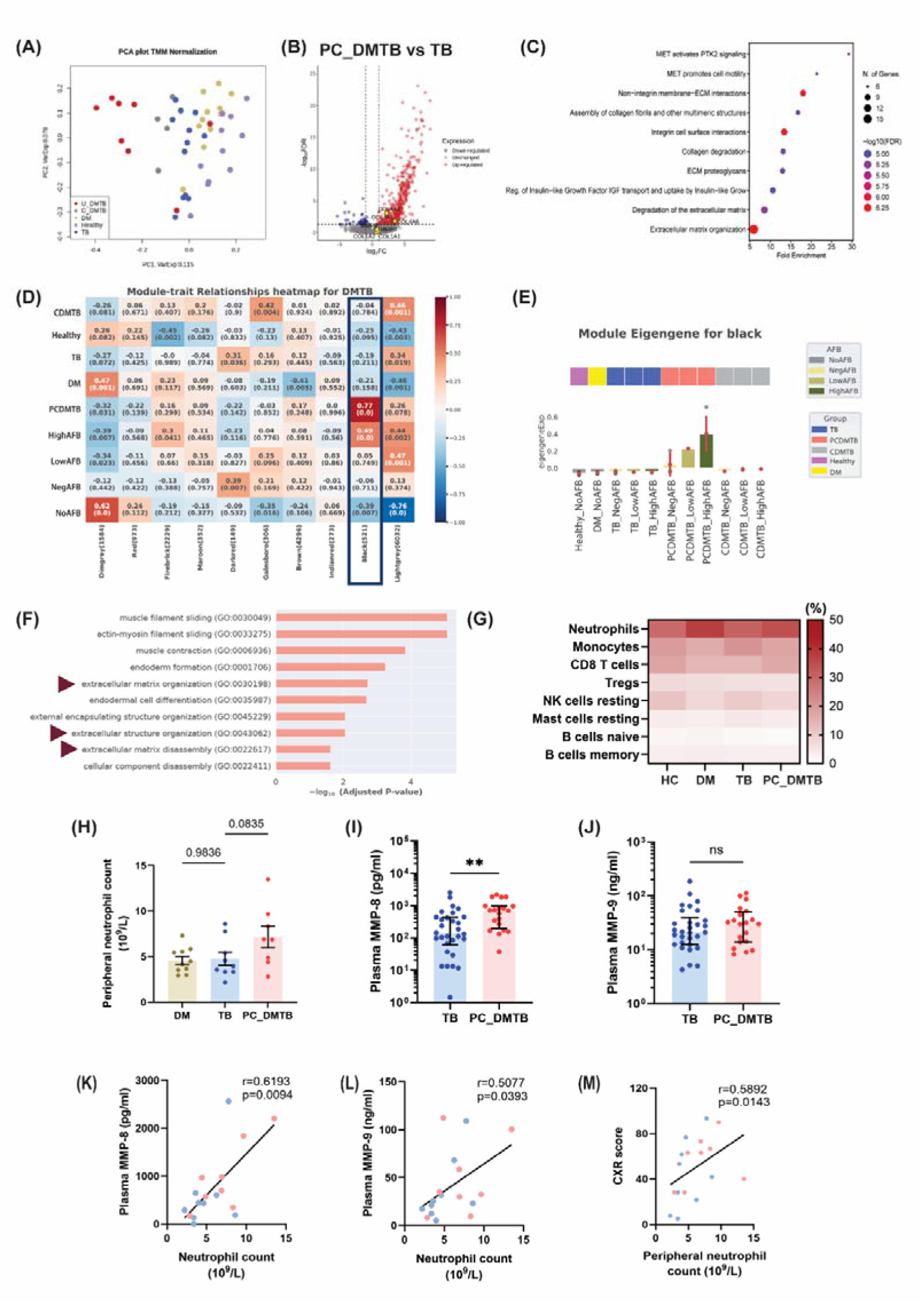
Whole blood RNA sequencing demonstrated increased neutrophil fraction in poorly-controlled diabetic TB patients with functional enrichment in extracellular matrix organisation. (A) PCA plot analysis of 11 HC, 11 DM patients, 12 TB patients, 4 C_DMTB patients and 8 PC_DMTB patients. (B) Volcano plots showing distribution of gene expression fold changes for PC_DMTB relative to TB, with cut-offs of FDR<0.05 and log_2_ FC larger than absolute 1. Red, upregulated genes; Blue, downregulated genes. (C) ShinyGO analysis of Reactome database revealed changes in pathways associated with collagen degradation, extracellular matrix degradation and extracellular matrix organisation in poorly-controlled DMTB patients. (D-F) Weighted gene co-expression network analysis. (D) Module-trait relationship analysis identified strong association (r=0.77) between black module and poorly-controlled DMTB patients. (E) Black module eigengene expression profile categorised by sputum AFB smear burden. The overall expression increased with increased sputum AFB smear burden in poorly-controlled DMTB patients. Top row, patient groups; bottom bar plots, module eigengene expression by AFB burden. NoAFB: no AFB test performed; NegAFB: AFB smear score 0; LowAFB: AFB smear score 1–2; HighAFB: AFB smear score 3–4. (F) GO enrichment analysis identified genes in black module associated with ECM organisation and disassembly. (G) Cibersortx predicts an increase in neutrophil cell population in peripheral blood from DM and poorly-controlled DMTB patients relative to HC. (H) Trend of higher peripheral neutrophil count in poorly-controlled DMTB patients compared to non-diabetic TB patients. Data are presented as mean ± SEM. One-way ANOVA test with Dunnet’s multiple comparison test was performed. n=10 DM, n=9 TB, n=8 PC_DMTB (I) Higher plasma MMP-8 concentrations in poorly-controlled DM-TB patients relative to TB, while (J) no difference was observed for plasma MMP-9. n=30 TB, n=20 poorly-controlled DMTB patients. Data are presented as median and interquartile range. Mann-Whiney test was performed. (K & L) Positive correlation between plasma MMP-8 and MMP-9 with peripheral neutrophil count. (M) Positive correlation between peripheral neutrophil count and CXR score. \**p*<0.05, \*\**p*<0.01.

Using Cibersortx (29), higher circulating neutrophil proportions was predicted in poorly-controlled DMTB (mean =34.1%) compared to TB (30.6%) and HC (28.8%) (**Figure 2G**). This observation was also consistent with the increasing peripheral neutrophil counts in poorly-controlled DMTB (Mean: non-diabetic TB 4.762 × 10L/L vs poorly-controlled DMTB 7.171× 10L/L, p=0.0835, **Figure 2H**). Plasma MMP-8 concentration was significantly up-regulated in poorly-controlled DMTB (median: non diabetic TB 138.7pg/mL vs poorly-controlled DMTB 707.5pg/mL, *p<*0.01, **Figure 2I**), while MMP-9 remained unchanged (**Figure 2J**). Both MMP-8 and MMP-9 were associated with peripheral neutrophil counts (**Figure 2K-L**), and neutrophilia was associated with worse CXR scores (**Figure 2M**), reinforcing neutrophil-driving increased MMP activity and lung damage. Systemically, other MMPs did not differ between poorly-controlled DMTB and non-diabetic TB, whereas TIMP-2 was further reduced in poorly-controlled DMTB, in keeping with the respiratory finding (**Figure S3**). Taken together, these findings suggest neutrophils may be implicated in ECM destruction in DMTB pathogenesis.

### Heterogenous neutrophil subset changes are found in DMTB patients at the single-cell level

To further determine the neutrophil gene expression changes, purified human primary neutrophils from six HC, four DM, five TB and five DMTB (two controlled-DMTB and three poorly-controlled DMTB) patients were subjected to scRNA sequencing. Uniform manifold and projection (UMAP) identified nine neutrophil subsets (**Figure 3A**), annotated based on established marker genes (33): cycling neutrophils (high *MKI67* and *PCNA*), myelocytes (high *LTF*, *LCN2* and *CAMP*), immature, maturing and mature neutrophils (high, intermediate and low expression of *PADI4* and *MME* respectively), maturing (interferon) neutrophils (enriched for interferon-stimulated genes), interferon-responsive neutrophils (high interferon-stimulated genes such as *MX1*, *IFIT2*, *STAT1*, *GBP1* and *GBP5)*, aged neutrophils (*CXCR4* and *TNFAIP2)* and long-lived neutrophils (high *PI3* and *SIGLEC10)* (**Figure 3A-B**). The top and representative genes defining each subset are shown in **Figure 3B**.

**Figure 3.**
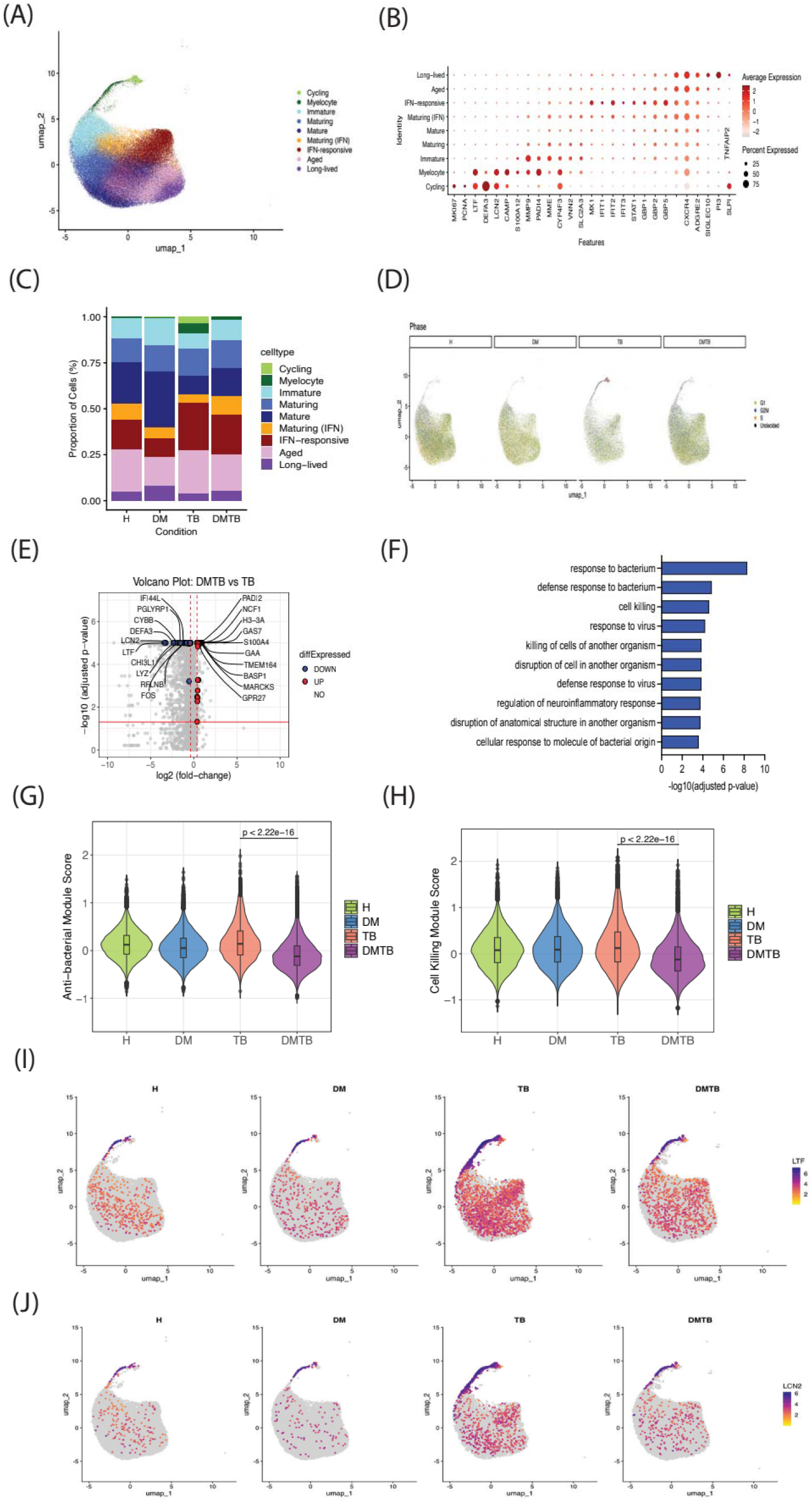
Reduced cell killing and anti-bacterial response in DMTB neutrophils at single-cell level. (A) UMAP showing 38,882 cells from HC (n=6), 26,789 cells from DM (n=4), 18,036 cells from TB (n=5) and 35,118 cells from DMTB (n=5). Nine neutrophil clusters were identified. (B) Dot plot showing expression of marker genes used for clusters annotation. (C) Proportion of cell types across 4 conditions: HC, DM, TB and DMTB. (D) UMAP showing cell cycle scoring across 4 conditions: HC, DM, TB and DMTB. (E) Volcano plot showing differentially expressed genes (DEGs) expressions in DMTB versus TB patients. The expression difference is considered significant for fold-change more than or less than 1.3 (vertical dashed red lines), adjusted p-value less than 0.05 (horizontal solid red line) and expression is seen in at least 10% of cells in either conditions. (F) Pathway enrichment analysis of the genes differentially expressed in DMTB compared to TB as in (E) performed using enrichGO. The bar chart displays the top 10 most significant pathways, considering both up– and down-regulated pathways, ranked by absolute enrichment p-value. (G–H) Violin plots showing pathway cell module scores (Seurat’s AddModuleScore) for (G) anti-bacterial response pathways and (H) cell killing pathways identified in (F) across 4 conditions: HC, DM, TB and DMTB. Statistical significance was assessed using the Wilcoxon rank-sum test. (I-J) UMAP showing the expression of granules-related genes LCN2 (lipocalin 2 or NGAL) and LYZ (lysozyme) across 4 conditions: HC, DM, TB and DMTB. Both genes are components of the antibacterial response and cell killing pathways in F-H.

The ratio of each neutrophil subset across the conditions was then calculated. The proportion of cycling neutrophils and myelocytes was higher in TB patients compared to DMTB, DM and HC (**Figure 3C and S4**). The cycling neutrophils exhibited enhanced proliferative signatures, with increased expression of G2/M and S phase markers (**Figure 3D and S5**). These findings suggest expansion of neutrophil precursors in healthy controls who acquired TB, but not in DMTB patients.

Differential expression analysis of total neutrophils between DMTB and TB patients revealed more significantly downregulated genes compared to upregulated genes in DMTB patients (**Figure 3E**). To further characterise these alterations, we performed pathway analysis of the differentially expressed genes (DEGs). The most significant pathways that were downregulated in DMTB patients were pathways associated with bacterial response and cell killing, indicating reduced antimicrobial function in DMTB patients (**Figure 3F**, **Supplementary file 2**). Consistently, the aggregate gene expression levels of DEGs involved in bacterial response and cell killing was also significantly reduced in DMTB compared to TB patients (**Figure 3G-H**). Notably, *LCN2* (lipocalin 2 or NGAL) and *LTF* (lactoferrin) were among the top DEGs (**Figure 3E**), and were predominantly expressed in cycling neutrophils and myelocytes (**Figure 3I-J, Figure S6**). Their expression was significantly higher in myelocytes from TB patients compared to DMTB patients, suggesting reduced granule-synthesis leading to weaker degranulation-dependent killing in DMTB (**Figure S6)**. Collectively, these data indicate that DMTB patients exhibited reduced granule-related genes in myelocytes with an overall reduced expression of bacterial response and cell killing genes that may have exacerbated TB pathogenesis.

### Altered neutrophil responses to mycobacteria are found in poorly-controlled DM

Given the pronounced tissue-destructive phenotype in poorly controlled DMTB patients, together with respiratory and systemic data implicating neutrophils in inflammatory and tissue-destructive responses, we isolated peripheral blood neutrophils from HC and DM patients to further investigate their functional responses following *Mtb* stimulation. Compared to HC, DM neutrophils produced significantly higher intracellular and extracellular ROS following *Mtb* stimulation (**Figures 4A-B**). Neutrophil extracellular trap (NET) formation, a process involving the release of decondensed chromatin and antimicrobial proteins to trap pathogens (40), showed a reduced trend, with suppressed extracellular DNA and neutrophil elastase (NE)-DNA complex in *Mtb*-stimulated DM neutrophils (**Figure 4C**). Plasma neutrophil elastase, another antimicrobial neutrophil peptide found on NETs, was 0.78-fold lower in poorly-controlled DMTB patients (*p*<0.05, **Figure 4D**). DM neutrophils also exhibited delayed phagocytic uptake of mycobacteria at 10 minutes (23.5% ± 2.43% in DM vs. 38.9% ± 5.43% in HC, **Figure 4E**). While MMP-8 remained unchanged on *Mtb* stimulation, DM neutrophils released 1.24-fold more MMP-9 than HC (*p*<0.01, **Figure 4F**). *Mtb*-stimulated DM neutrophils also showed up-regulated type I collagen-destructive activity (**Figure 4G**). Taken together, our results suggested that poorly-controlled DM patients have dysfunctional neutrophil responses following *Mtb* stimulation, which may drive increased TB severity in DMTB patients.

**Figure 4.**
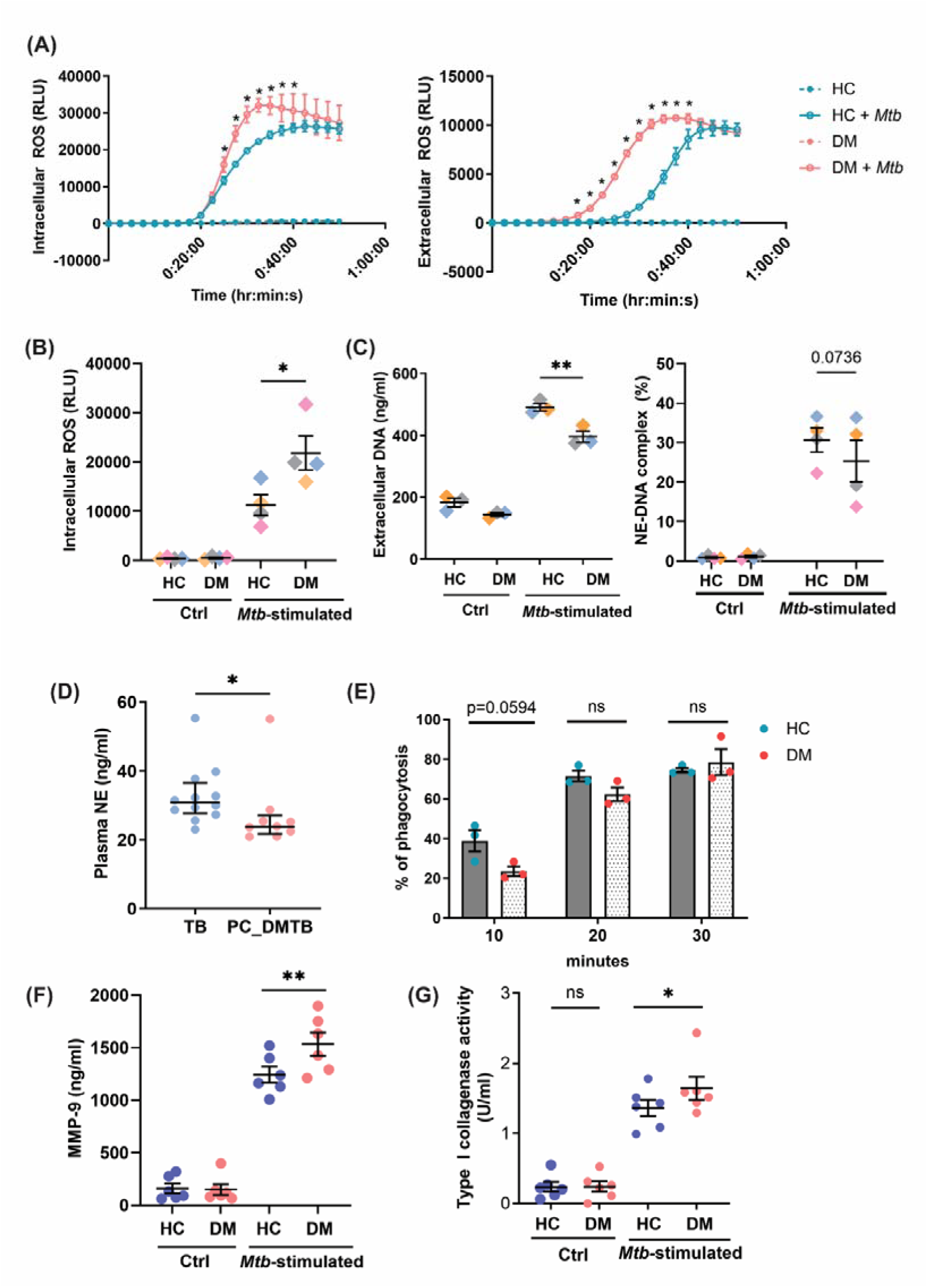
Neutrophil dysfunction to mycobacteria stimulation is found in poorly-controlled DM. (A) Neutrophil intracellular and extracellular ROS production over time. Data shown are representative of 4 and 2 independent neutrophil donor pairs (HC+DM = 1 pair) respectively, with bars representing mean ± SD of technical triplicates. (B) Intracellular ROS production at 25^th^ minute of live *Mtb* stimulation. Data are presented as mean ± SEM from 4 independent donors from each set. Experiment was conducted in pairs with each colour representing one experimental set. (C) Neutrophil extracellular trap was assessed by measuring the amount of extracellular DNA and neutrophil elastase (NE)-DNA complex in the neutrophil supernatant using Picogreen and ELISA respectively. (D) Plasma NE concentration. n=12 TB, n=9 poorly-controlled DMTB. Mann-Whitney test was performed. (E) Percentage of neutrophils that have phagocytosed m-cherry labelled *M. bovis* BCG was determined by flow cytometry. Data are presented as mean ± SEM from 3 independent donors from each group. (F) Neutrophil MMP-9 released is increased in DM neutrophils. (G) Type I collagenase activity was upregulated in DM-neutrophils following *Mtb* stimulation. (A), (B), (C), (E), (F) and (G) Two-way RM ANOVA and Sidak’s multiple comparison test was conducted. *Adjusted *p*-value<0.05, **Adjusted *p*-value<0.01.

### Increased TNFR and RAGE expression in poorly-controlled DM neutrophils after *Mtb*-stimulation

We sought to determine signalling pathways that may drive neutrophil hyperinflammatory responses. Activation of NF-kB signalling induces pro-inflammatory mediators’ gene expression and protein production (41, 42). In neutrophils, NF-kB activation promotes NETs (43), MMP-8 and MMP-9 release (23, 44), all of which contribute to TB immunopathology (14, 23). To explore the contribution of NF-κB pathway to neutrophil-driven inflammation in DMTB, we performed an NF-κB array which revealed upregulation of TNFR1 and TNFR2 in DM neutrophils following 30-minute *Mtb* stimulation (**Figures 5A-B**) and further confirmed by western blot (**Figures 5C-D**). Basal levels remained unchanged in healthy and diabetic unstimulated neutrophils (**Figure S7A-B**). Neutrophil TNF-α and soluble TNFR1 and TNFR2 also remained unchanged following *Mtb* stimulation (**Figure S7C-E**). Poorly-controlled DMTB patients showed increasing trend of higher plasma TNF-α and soluble TNFR2 (sTNFR2) (**Figures 5E-F**), and whole blood transcriptomics indicated TNF-α pathway enrichment (**Figure 5H**). This was mirrored in the respiratory compartment, where upregulated sputum TNF-α, TNFR1, and TNFR2 (**Figure 5I-K**) implicated TNF-driven hyperinflammatory signalling in poorly-controlled DMTB patients.

**Figure 5.**
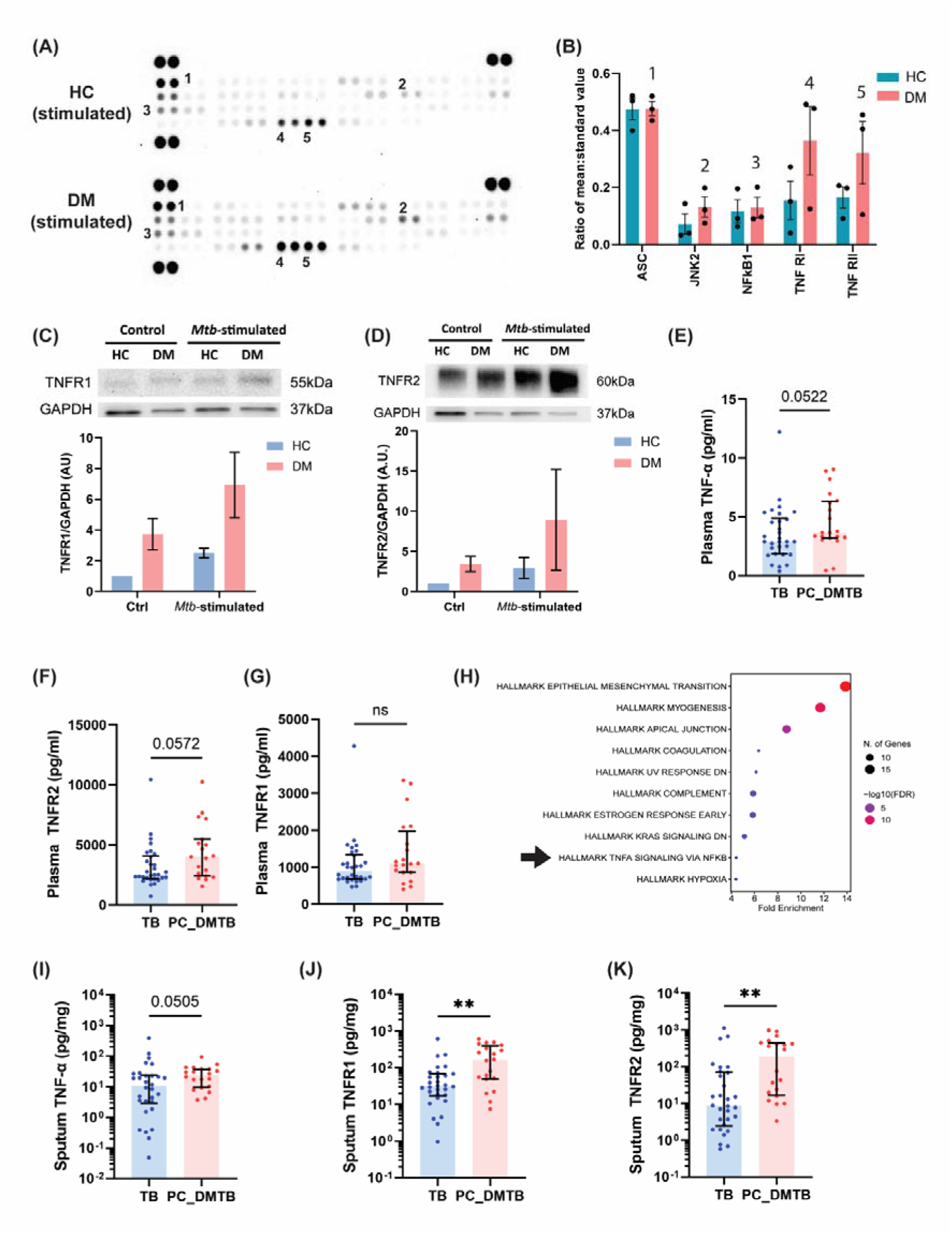
Increased TNFR1 and TNFR2 activation drive hyperinflammation in poorly-controlled DMTB disease. (A) NF-κB array analysis of neutrophils from HC and poorly-controlled DM patients following *Mtb* stimulation showed increased TNFR1 and TNFR2 protein expression in DM neutrophils. (B) Densitometric analysis was performed using FIJI. Data are presented as mean ± SEM, from 3 independent donors in each set. Western blot analysis showed increased (C) TNFR1 and (D) TNFR2 proteins in live *Mtb* stimulated DM neutrophils compared to HC. Infection experiment was performed in pairs, consisting of 1 HC and 1 DM patient each time, and the experiment was performed twice in two different pairs of donors. Trend towards upregulated plasma (E) TNF-α and (F) TNFR2 levels were observed in poorly-controlled DMTB. (G) Plasma TNFR1 remained unchanged between the two groups. (H) Gene functional enrichment analysis showed enrichment in pathway associated with TNF-α signalling in TB patients with poorly-controlled DM, in keeping with the cellular model findings. Analysis was performed using hallmark database and differentially upregulated genes from TB patients with poorly-controlled DM relative to TB patients without DM, with false discovery rate (FDR)<0.05. Sputum concentrations of (I) TNF-α, (J) TNFR1, and (K) TNFR2 were upregulated in poorly-controlled DMTB patients.

As diabetes is associated with enhanced AGE–RAGE signalling and increased tissue inflammation, and given the elevated circulating RAGE ligands in DMTB patients (45–47), we next evaluated RAGE expression. RAGE activity is regulated by metalloproteinase-dependent shedding and alternative splicing, generating soluble RAGE (sRAGE), which can act as a decoy receptor to bind circulating ligands and limit RAGE-mediated pro-inflammatory signalling (48, 49). We found *Mtb*-stimulated DM neutrophils expressed higher levels of full-length RAGE compared to HC (**Figure 6A**). In contrast, plasma concentrations of anti-inflammatory sRAGE were significantly suppressed in poorly-controlled DM and poorly-controlled DMTB patients compared to HC and TB patients respectively (**Figures 6B–C**). Plasma RAGE ligand, S100A12 was markedly increased in poorly-controlled DMTB patients (**Figure 6D**), and was further confirmed by KEGG analysis, which identified enrichment in AGE-RAGE pathway (**Figure 6E**). Transcriptional upregulation of RAGE ligands (S100A8, S100A9, S100A12) was also observed (**Figure 6F**). As RAGE may be further mitigated using existing anti-diabetic drugs, we determined if RAGE could be modulated using rosiglitazone. Following rosiglitazone treatment, we found reduced ROS and MMP-8 secretion following *Mtb* stimulation in DM neutrophils (**Figures 6G–H),** highlighting the therapeutic potential of targeting RAGE signalling in DMTB pathology. Together, these findings indicate dysregulated AGE/RAGE, concurrently with hyper-inflammatory TNF signalling in DMTB, amplifies neutrophil-driven hyperinflammation. Pharmacological modulation of these pathways may help mitigate immunopathology and improve clinical outcomes in DMTB patients.

**Figure 6.**
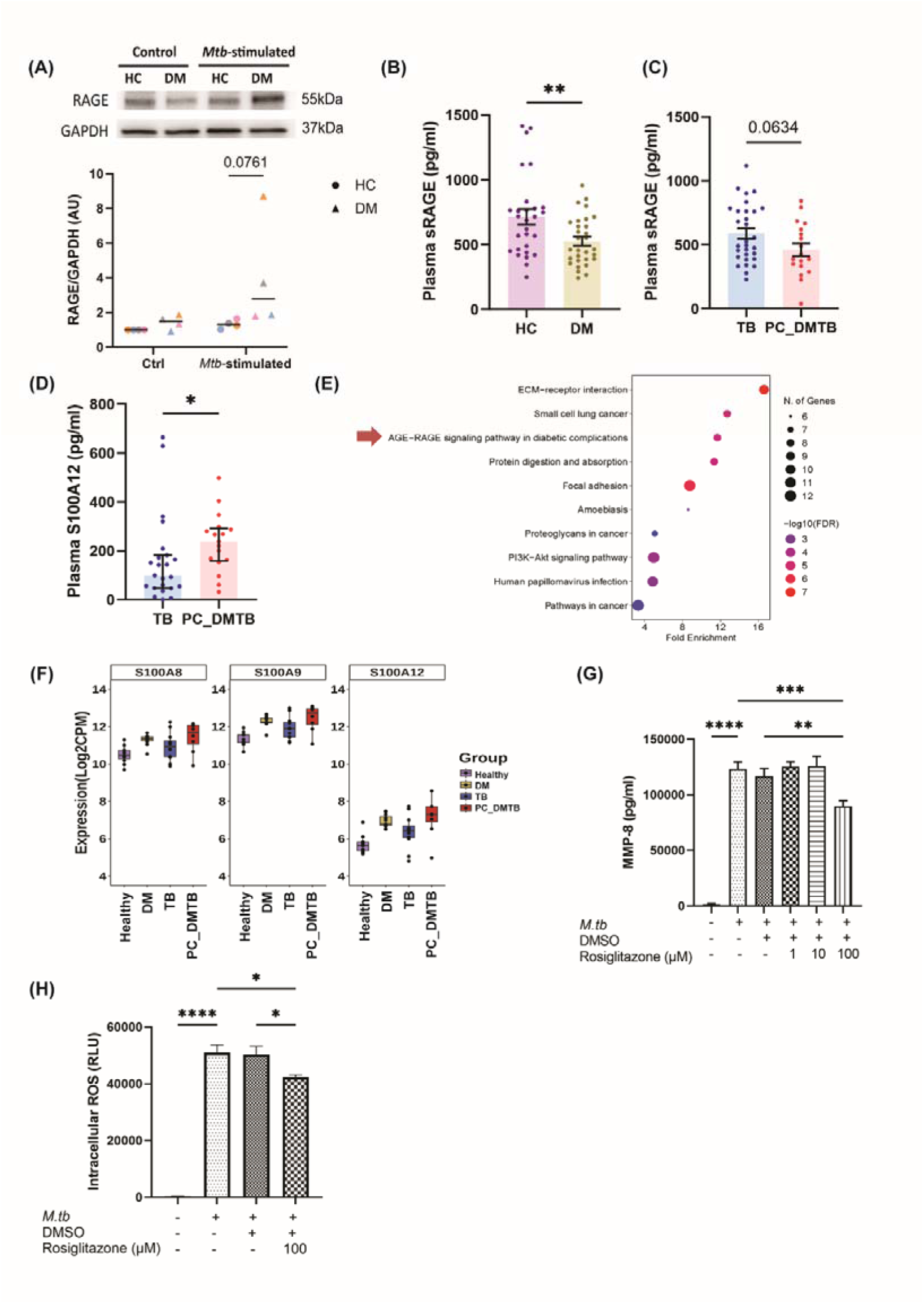
Increased RAGE activation drives hyperinflammatory response in PC_DMTB. (A) Western blot analysis showed increased full length RAGE protein expression in *Mtb-*stimulated neutrophils from DM patients. Representative blot was shown. Bars represent the median from n=4 donors from each group. Two-way ANOVA test with Sidak’s multiple comparison test was performed. (B) DM patients had lower anti-inflammatory soluble RAGE in the plasma. n=28 HC and n=30 DM. (C) Poorly-controlled DMTB patients had lower plasma soluble RAGE compared to non-DM TB patients. n=30 TB, n=18 PC_DMTB. (D) Plasma inflammatory RAGE ligand S100A12 was higher in poorly-controlled DMTB patients. n=23 TB, n=17 PC_DMTB. (E) KEGG pathway analysis revealed that the upregulated DEGs in TB patients with poorly-controlled DM were associated with AGE-RAGE signalling, indicating perturbation of AGE/RAGE pathway in poorly-controlled DMTB. (F) RAGE ligand gene expression of S100A8, S100A9 and S100A12 were relatively higher in PC_DMTB patients compared to non-DM TB patients. n=11 HC, n=11 DM, n=12 TB, n=8 PC_DMTB. RAGE inhibition by rosiglitazone treatment suppressed (G) MMP-8 and (H) ROS release from *Mtb-*stimulated neutrophils. Data shown are representative of at least 2 donors. Two-ways ANOVA test with Sidak’s multiple comparison test were performed. \**p*<0.05, \*\**p*<0.01, \*\*\**p*<0.001, **** *p*<0.0001.

## Discussion

Poorly-controlled DM worsens pulmonary TB pathology, but mechanisms remain unclear. While previous studies have largely focused on impaired adaptive immunity in DMTB (50–53), our findings highlight a key role for neutrophil-driven inflammation across both respiratory and systemic compartments. In the respiratory compartment, respiratory secretions from poorly controlled DMTB patients showed markedly elevated neutrophil-associated MMP-8 and MMP-9, accompanied by increased matrix-destruction. These changes were associated with neutrophil markers and radiographic evidence of lung damage, including cavitation, implicating neutrophil-driven proteolysis in pulmonary tissue destruction.

Systemically, whole blood transcriptomic analysis revealed enrichment of extracellular matrix degradation pathways and increased predicted neutrophil fractions in poorly-controlled DMTB, alongside elevated circulating MMP-8, indicating coordinated activation of neutrophil-associated inflammatory and proteolytic programs across systemic and respiratory compartments. The systemic MMP findings differ from those of Kumar *et al*., who reported increases in other MMPs but not MMP-8, and observed metformin-associated reductions in MMP levels that were not replicated in our cohort (19). These discrepancies may reflect differences in cohort characteristics, including glycaemic control, HbA1c distribution, and ethnicity or sample collection differences.

At single-cell resolution, neutrophils from DMTB patients exhibited reduced module scores for cell-killing and bacterial-response pathways, suggesting impaired antibacterial programming that may contribute to higher sputum mycobacterial burden. LYZ, DEFA3, LCN2 and LTF which encode the granule proteins lysozyme, defensin alpha 3, lipocalin-2 (NGAL) and lactoferrin, respectively, were downregulated. As these genes are enriched in cell-killing and bacterial-response pathways, their reduced expression indicates a defect in granule-mediated antimicrobial activity (54). This is supported by prior studies linking hyperglycaemia to impaired neutrophil degranulation, providing a potential mechanism for poorer outcomes in DMTB (55). Neutrophils precursors typically express high levels of granule proteins transcripts, which decline upon maturation (56). In our datasets, the cycling population and myelocytes likely represented the precursor populations with elevated expression granule genes such as LCN2 and LTF. Cycling neutrophils appeared proliferative – an uncommon feature in circulating neutrophils. Myelocytes, in contrast, were non-proliferating precursors but lower granule expression in DMTB compared to TB suggested weaker degranulation-dependent killing. Both subsets may correspond to low density granulocytes (LDGs), which besides being more buoyant in density, were characterised by expression of granule proteins and cell-cycle genes transcripts (**Figure S8)** (57, 58). The characteristics and functions of LDGs remain contentious with conflicting findings in different disease context (58). In TB, the role of LDGs remains incompletely defined but their expansion has been linked to increased disease severity (59). The significance of the higher proportion of neutrophil precursors in TB compared with DMTB remains uncertain; however, it is plausible that robust anti-microbial activity involving these precursors in TB may improve containment of TB disease compared to DMTB. Definitive clarification on the roles of these neutrophil subsets will require complementary protein-level and functional studies.

In our cellular TB model, neutrophils from poorly-controlled DM patients exhibited a dysregulated phenotype upon mycobacterial stimulation, characterised by increased MMP-9 and ROS production, impaired NET formation, and delayed phagocytosis. While neutrophil dysfunction in diabetes has been described under non-TB or chemical stimulation conditions (60–65), our study extends these observations to *Mtb-*driven responses. Although MMP-8 release was not significantly different between groups, type I collagenase activity was increased, indicating enhanced proteolytic activity likely driven by active MMPs and/or additional proteases. Elevated ROS production in DM neutrophils may reflect hyperglycaemia-induced metabolic reprogramming via polyol pathway flux, nicotinamide adenine dinucleotide phosphate (NADPH) depletion limiting glutathione regeneration, and enhanced mitochondrial ROS generation (66). While prior studies using phorbol 12-myristate 13-acetate reported reduced ROS in DMTB (67), our use of live *Mtb* elicited robust ROS production, highlighting stimulus-specific responses. Reduced NET formation contrasts with chemical stimulation studies (60, 61, 63), but aligns with bacterial exposure model in poorly controlled DM (65). Similarly, delayed phagocytic uptake is consistent with prior reports of impaired phagocytosis in diabetes (64). Functionally, these defects likely impair bacterial containment, consistent with a trend toward increased sputum AFB burden in our cohort and others of impaired mycobacterial clearance in DMTB (68).

Mechanistically, TNF signalling was enriched in poorly-controlled DMTB. While TNF signalling is well described in TB (69–71), its regulation in neutrophils during *Mtb* infection in poorly-controlled DM remains unclear. Here, *Mtb*-stimulated DM neutrophils showed increased surface expression of TNFR1 and TNFR2, despite similar levels of TNF-α, sTNFR1, and sTNFR2 in culture supernatant. Elevated circulating TNF-α *in vivo* may prime neutrophils for this response (72), consistent with higher plasma TNF-α in our DM patients. Poorly-controlled DMTB patients also had higher circulating soluble TNFRs in plasma and sputum. However, neutrophils did not show increased shedding *in vitro*, suggesting contributions from other immune and endothelial cells (73, 74). Functionally, increased TNFR expression may enhance NF-κB signalling and NADPH oxidase 2 (NOX2)-mediated ROS production, contributing to intracellular oxidative stress observed in DM neutrophils (75, 76).

RAGE signalling also contributes to neutrophil-driven inflammation. Elevated RAGE ligands have been reported in DMTB plasma (45), and RAGE is upregulated in diabetes and highly expressed in lung epithelium [127, 348]. However, the role of RAGE signalling in neutrophils during *Mtb* infection in the context of poorly-controlled diabetes has not been well defined. In our study, *Mtb-*stimulated DM neutrophils exhibited increased full-length RAGE expression compared to HC, while sRAGE, an anti-inflammatory decoy receptor, was reduced in poorly-controlled DMTB plasma compared to TB. In parallel, upregulation of RAGE ligand genes (S100A8/A9/A12) and elevated plasma S100A12 supported enhanced RAGE activity in poorly-controlled DMTB. These findings suggest a proinflammatory loop in which increased ligand availability and reduced sRAGE sustained NF-κB activation and ROS production. Inhibition of RAGE with rosiglitazone, a peroxisome proliferator-activated receptor gamma (PPARγ) agonist, significantly suppressed *Mtb*-induced ROS and MMP-8 release in neutrophils, consistent with prior reports of RAGE blockade reducing lung inflammation in various disease models (16, 77–80). Collectively, these results highlight RAGE as a potential therapeutic target to mitigate neutrophil-mediated tissue damage in poorly-controlled DMTB.

This study has several limitations. The cross-sectional design limits causal inference and precludes assessment of temporal dynamics in neutrophil dysfunction during disease progression and treatment. The lack of experimental validation in animal models limits confirmation of mechanistic pathways identified in human samples. Future studies incorporating longitudinal sampling and animal models are needed to validate these findings and further define neutrophil dysregulation in DMTB, with a view toward developing targeted host-directed therapies.

Altogether, our study supports a prominent role for neutrophils in lung destruction and impaired bacterial control in TB under poorly-controlled DM. scRNA-sequencing revealed neutrophil subsets with altered antimicrobial programming, while *in vitro* assays showed increased ROS, altered MMP activity, delayed phagocytosis, and reduced NET formation. Dysregulation of TNF and RAGE signalling may drive excessive inflammation and tissue injury in poorly-controlled DMTB. Targeting RAGE alongside tight glycaemic control would mitigate neutrophil-mediated immunopathology to improve outcomes.

## Data availability

We deposited the bulk RNA-seq data in the Gene Expression Omnibus (GEO) repository (GSE328534) and the scRNA-seq data in ArrayExpress (E-MTAB-XXXXX).

## Acknowledgement

We thank the team at the National University Hospital Investigational Medicine Unit and the Singapore Infectious Diseases Clinical Research Network for their assistance with patient recruitment and sample collection. We also acknowledge the National University of Singapore (NUS) Metabolic Core Facility for providing phlebotomy services for healthy donors. We are grateful to the operations team of the NUS BSL-3 Core Facility for their logistical support. Finally, we thank the NUS Medicine Flow Cytometry Laboratory Unit for providing access to equipment and for their valuable technical advice on the flow cytometry experiments.

## Author contributions

C.W.M.O. conceived the study and obtained funding. P.M.T., T.H.H., F.K.L., H.L., C.B., H.T.C., and A.J.W.C. conducted the experiments. C.W.M.O., K.R.C., P.M.T., T.H.H., J.S.G.O., F.K.L., and A.F.V. analysed the data. A.F.V., F.K.L., and P.M.T. contributed to whole-blood RNA-seq analysis. T.H.H., J.S.G.O., and K.R.C. carried out bioinformatic analysis of single-cell transcriptomic data. C.V.C., L.C.G., L.H.W.L., S.P.Y., A.Y.L.L., S.F.M., S.L.K., and C.W.M.O. screened and recruited study participants. A.X.Y.A., D.L.L.B. and C.W.M.O. scored the chest X-ray images. P.M.T., T.H.H., K.R.C., and C.W.M.O. wrote the first draft of the paper which was reviewed and significantly revised by all authors.

