## Supplemental materials for "Neutrophil-primed immunopathology in poorly-controlled diabetes worsens matrix destruction in pulmonary tuberculosis"

**Supplementary Files:**

1. Supp file 1_DMTB study protocol
2. Supp file 2_GOBP_ORA_DEG_sig

### Supplementary Method

#### 1. ELISA

Quantification of S100A8/S100A9 heterodimer, S100A12, receptor for advanced glycation end products (RAGE), and neutrophil elastase was performed using ELISA kits from R&D Systems (S100A8/A9: DY8226-05; S100A12: DY1052-05; RAGE: DRG00; elastase: DY9167-05), following the manufacturers' protocols.

#### 2. Type I collagen and elastase degradation assays

Type I collagen and elastin degradation were evaluated using EnzChek® Collagenase (#E12055) and Elastase (#E12056) assay kits (Invitrogen). performed according to the manufacturer's instructions. Neutrophil supernatants were diluted 1:5 in 1× reaction buffer. Induced sputum samples were diluted 1:5 or 1:50 for collagenase assays and 1:2 or 1:20 for elastase assays. Samples were activated with 4-amino-phenyl mercuric acetate (APMA) at final concentration of 2mM for 1 hour at 37℃. Standard curves were prepared using 2-fold serial dilutions of enzyme standards ranging from 1 U/mL to 0.0156 U/mL. For collagenase, 40µl 1x reaction buffer was added into each well, followed by 10µl DQ collagen at final working concentration of 100µg/ml, and 50µl activated samples or standards. For elastase assay, 25µl 1x reaction buffer, 25µ DQ elastin at final working concentration of 25µg/ml, and 50µl activated samples or standards were added. The samples were incubated at room temperature in the dark, and fluorescence was measured at specified times using Synergy H1 microplate reader (BioTek, USA).

#### 3. Whole blood bulk RNA sequencing

##### 3.1. RNA extraction and library preparation

Blood samples collected into Tempus tube were processed as per manufacturer's protocol to obtain the RNA pellet. The pellet was dissolved in TRIzol (Thermo Fisher Scientific, #15596026). Total RNA was extracted by acid guanidinium thiocyanate-phenol-chloroform extraction and cleaned up using Qiagen RNeasy kit. The mRNA was enriched using poly (A) capture method and reverse transcribed to cDNA. The cDNA library generated is sequenced on Illumina NovaSeq platform, using 150bp paired-end sequencing strategy.

#### 3.2. Differential gene expression (DEG) analysis

Raw counts from RNA sequencing were processed using Bioconductor package EdgeR (v3.17)(1), with 3 variances estimated and size factor normalized using Trimmed Mean of M-values (TMM). Differential analysis was performed using EdgeR and filterByExpr function was used to filter genes that have low number of reads. Samples with a median exceeding twice the median absolute deviation (MAD) from the median of medians, as well as observations more than two IQR below Q1 or above Q3, were considered outliers and excluded from downstream analysis. Genes with a false discovery rate (FDR)-corrected p-value<0.05 are considered differentially expressed, following on from a likelihood ratio test using a negative binomial generalized linear model fit. Limma with its voomWithQualityWeights function (version 3.56.2) was performed and the counts were used for principal component analysis (PCA)(2).

#### 3.3. Gene enrichment analysis

ShinyGO v0.77 online tool (http://bioinformatics.sdstate.edu/go/) (3) was used for gene functional enrichment analysis. The list of differentially upregulated genes from patients with false discovery rate (FDR)<0.05 was submitted for analysis. The Kyoto Encyclopedia of Genes and Genomes (KEGG), Gene Ontology Biological Processes (GOBP) and Reactome databases were used to explore the potential function of the differentially upregulated genes.

#### 3.4. Immune cell fraction estimation using Cibersortx

Cibersortx (4, 5) was used to estimate immune cell fractions in whole blood without physical isolation, utilising the LM22 signature matrix of 547 genes to distinguish 22 immune cell subtypes (6). A B-mode batch correction was applied to account for platform differences between the microarray-derived LM22 matrix and bulk RNA-sequencing data, while quantile normalization was disabled (4, 7, 8).

#### 3.5. Weighted gene co-expression network analysis (WGCNA)

PyWGCNA is a python library designed to perform WGCNA analysis. WGCNA is a tool used to identify clusters (or modules) of highly correlated genes and examine their relationship with clinical traits(9). The analysis was run as described (10).

### 4. Single-cell RNA sequencing

#### 4.1. Neutrophil isolation and fixation

Untouched neutrophils were isolated from EDTA-anticoagulated whole blood using the EasySep™ Human Neutrophil Isolation Kit. Immediately after isolation, neutrophils were fixed and stored according to the 10x Genomics demonstrated protocol “Fixation of Cells & Nuclei for Chromium Fixed RNA Profiling” (Document CG000478, Rev D) for peripheral blood mononuclear cells (PBMCs) with minor change: Isolated neutrophils were centrifuged at 250 rcf for 10 min at RT prior to adding Fixation buffer.

#### 4.2. Library construction

Fixed neutrophils were processed with Chromium Fixed RNA Profiling kit (10x Genomics, User Guide CG000527 Rev E) targeting ~10 000 cells per sample. All steps were performed exactly as described in the protocol; no additional clean-up cycles or reagent substitutions were introduced.

#### 4.3. BGI sequencing

Indexed libraries were pooled equimolarly and sequenced on a DNBSEQ-T7 instrument (BGI) using a 150 × 8 × 8 × 150 bp read layout, aiming for 3 × 10^4 paired reads per cell.

#### 4.4. FASTQ processing

FASTQ files were generated with bcl2fastq (BGI, default settings) and aligned with Cell Ranger v9.0.0 (10x Genomics) against the GRCh38-2024-A reference using default parameters.

#### 4.5. Low-quality cell and doublet filtering

Quality control criteria were: (1) total UMI count between 100 and 30,000, (2) minimal number of detected genes > 100, (3) mitochondrial gene percentage < 20 % and (4) number of cells expressing a gene > 10.

#### 4.6. Normalisation, HVG selection and dimensionality reduction

The following was conducted using Seurat (11). The count matrix was log-normalized and scaled. The top 29 dimensions from the principal component analysis (PCA) were used for the uniform manifold approximation and projection (UMAP). The top PCs were chosen based on the Elbow plot and Jackstraw methods. Sample integration was conducted using CCAIntegration.

#### 4.7. Clustering and annotation

Cell clusters were identified with the Leiden algorithm (resolution = 0.5) on the SCVI latent graph and annotated manually based on canonical neutrophil marker genes(12).

#### 4.8. Differential gene expression analysis

Differentially expressed genes between DMTB vs TB were determined using FindMarkers and using a cutoff of adjusted p-value < 0.05, min.pct > 0.1 for both conditions, FC cutoff of 1.3. Pathway analysis was conducted using enrichGO from the package clusterProfiler. Significant pathways were determined using a cutoff of adjust p-value < 0.05 and at least 5 or more genes per pathway.

### 5. *Ex vivo* culture

Neutrophils from HC and DM patients were stimulated in pairs with *Mtb* H37Rv at a multiplicity of infection (MOI) of 10 for 3 hours as described(13). For drug treatment experiment, neutrophils were preincubated with stated concentration of rosiglitazone (Sigma-Aldrich, #R2408) for 1 hour prior to *Mtb* stimulation.

### 6. Phagocytosis measured by flow cytometry

100µl lithium heparin blood was stimulated with m-Cherry labelled *M. bovis* BCG at MOI of 10 for 10, 20 and 30 minutes respectively. Neutrophils were stained with anti-CD66c marker (BD Biosciences, #742683), erythrocytes were lysed and cells were fixed using 4% formaldehyde solution (Sigma Aldrich) prior to analysis using BD LSR Fortessa flow cytometer. Neutrophils were identified based on forward and side scatter properties and CD66c expression. A minimum of 10,000 CD66c$^+$ events were acquired per sample. Phagocytosis was quantified as the percentage of CD66c$^+$ cells positive for mCherry signal. Data was analysed using FlowJo v10 (Flowjo, USA). Detailed gating strategy is shown in the figure below.

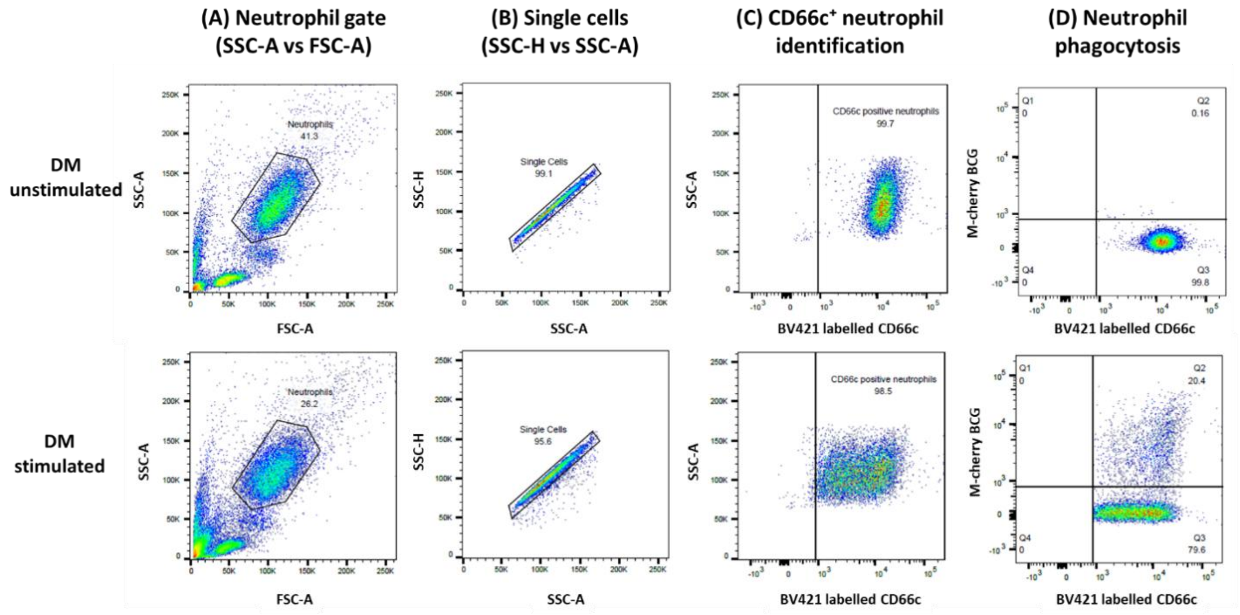

Gating strategy for neutrophil phagocytosis assay using whole blood. Representative plots from unstimulated and mCherry-labelled *Mycobacterium bovis* BCG-stimulated DM sample.
(A) Neutrophils were identified based on forward (FSC) and side (SSC) scatter properties.
(B) Single cells were selected using SSC-H versus SSC-A to exclude doublets.
(C) Neutrophils were confirmed by CD66c expression.
(D) Phagocytosis was quantified as the percentage of cells positive for uptake of mCherry-labelled *Mycobacterium bovis* BCG, with quadrant gating used to define phagocytosis-positive (Q2) and -negative (Q3) populations.

### 7. Neutrophil extracellular trap (NET) isolation and quantification

NETs were harvested by treating the cells with DNase I at final concentration of 0.6µg/ml for 15 minutes. EDTA was then added at a final concentration of 5mM to stop the reaction. The supernatant was harvested by spinning at 2000rpm for 1 minute and NETs were quantified using QuantiT Picogreen (Invitrogen, #P11496) and a neutrophil elastase-DNA ELISA.

For the ELISA, Nunc™ MaxiSorp™ 96-well plates were coated overnight at 4 °C with neutrophil elastase antibody (4 µg/mL; R&D Systems, DY9167-05). Plates were washed with PBST (PBS + 0.05% Tween 20) and blocked with 5% BSA (Lonza) for 2 hours at room temperature. NET standards were generated from pooled PMA-stimulated neutrophil supernatant from 5 healthy controls. Samples and standards (50 µl) were added in duplicate and incubated for 2 hours at room temperature with shaking. Supernatants from unstimulated neutrophils were diluted 1:5; *M. tuberculosis*–stimulated samples were diluted 1:50. After washing, 50µl of HRP-conjugated anti-dsDNA antibody (1:10 dilution; Roche, #11544675001) was added and incubated for 1 hour in the dark. Following four washes, 50 µl of TMB substrate (R&D Systems) was added for 20 minutes, and the reaction was stopped with 25µl of 2N sulfuric acid. The plate was read at 450nm and 540nm using Synergy H1 microplate reader (BioTek, USA).

### 8. Human NF-kB array

Human NF-kB array kit (R&D systems, #ARY029) is an antibody array that allows for simultaneous measurement of the relative levels of 41 NF-kB related human proteins and 4 phosphorylation sites. Neutrophil lysates (250 µg), collected 30 minutes after heat-killed *Mtb* stimulation, were analysed per the manufacturer's protocol. Densitometric analysis was done using FIJI.

### 9. Western blot

Stimulated neutrophils were pelleted and lysed in RIPA buffer (Thermo Scientific, #89900) containing complete proteases (Roche, #11873580001) and phosphatase inhibitors (Roche, #4906845001). Total protein concentration was determined using DC Protein Assay (Biorad) according to the manufacturer's instruction. Protein lysates were heat-denatured in 2× Laemmli sample buffer at 70 °C for 10 minutes and briefly cooled on ice. Equal amounts (20-25µg) of proteins were separated using 4-15% Mini-PROTEAN® TGX™ Precast Protein Gels (Biorad), and transferred onto PVDF membrane. Membranes were blocked and probed with primary antibodies against TNFR1, TNFR2 and RAGE protein, followed by chemiluminescent detection. Blots were imaged using the ChemiDoc Touch Imaging System (Bio-Rad) and analysed with Image Lab v6.0 software (Bio-Rad).

Primary and secondary antibodies used in western blot.

| Antibody | Manufacturer | Catalog number | Dilution |
|---|---|---|---|
| Primary Antibody | | | |
| Rabbit anti-human GAPDH | Cell Signaling Technology | #2118S | 1:5000 |
| Rabbit anti-human TNFR1 | Cell Signaling Technology | #3736S | 1:1000 |
| Rabbit anti-human TNFR2 | Cell Signaling Technology | #72337 | 1:1000 |
| Rabbit anti-human RAGE | Invitrogen | #PA5-24787 | 1:1000 |
| Secondary Antibody | | | |
| Goat anti-rabbit HRP linked | Cell Signaling Technology | #7074P2 | 1:1000 – 1:5000 |

### Supplementary Table

**Table S1 Spearman's correlation between sputum MMP concentration and type I collagenase activity**

| Analyte | Correlation coefficient, r | p-value |
|---|---|---|
| MMP-1 | **0.3391** | **0.0113** |
| MMP-8 | **0.7343** | **<0.0001** |
| MMP-13 | **-0.4212** | **0.0014** |

### Supplementary Figures

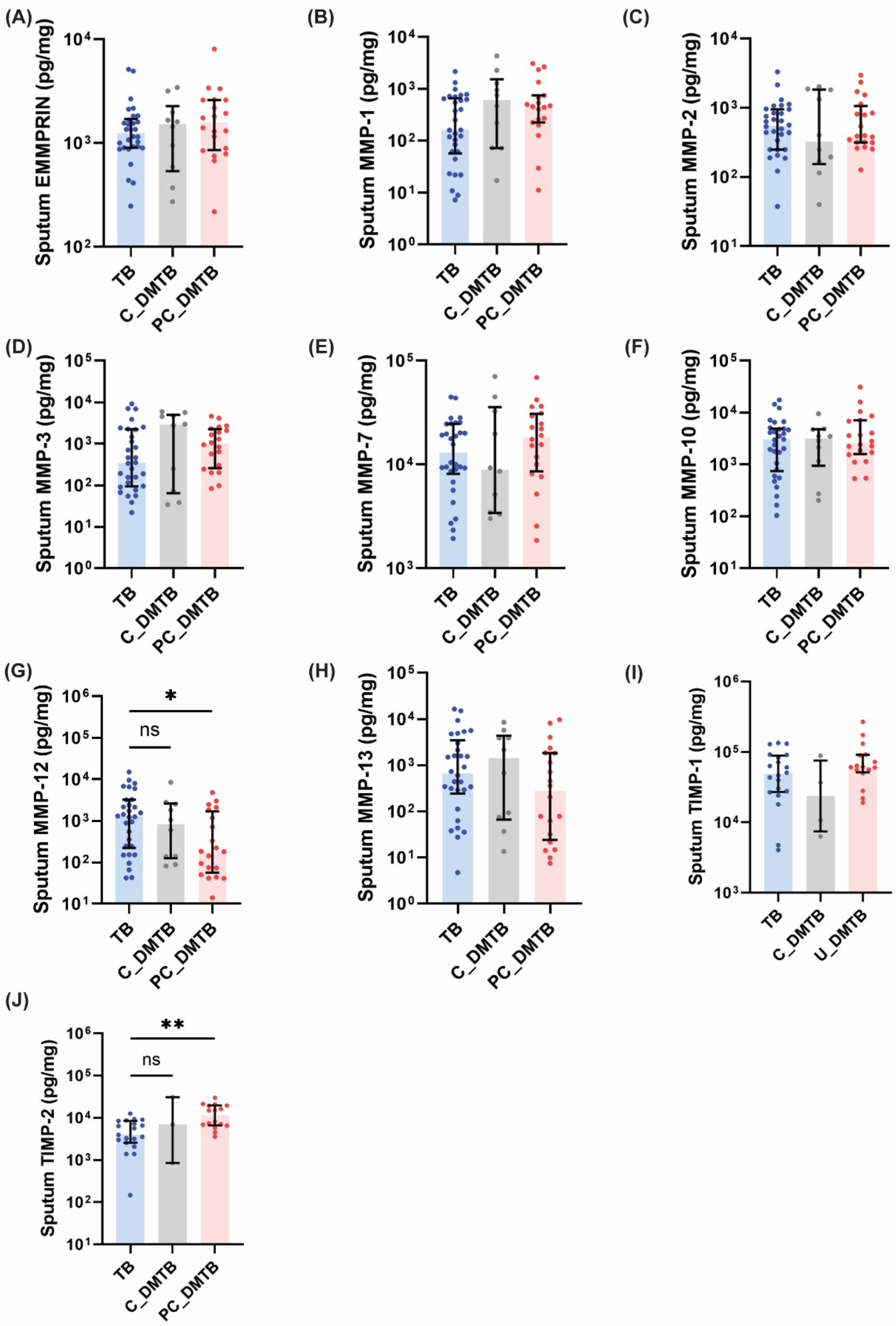

**Figure S1. Sputum EMMPRIN, MMP-1, -2, -3, -7, -10, -13, and TIMP-1/-2 in non-diabetic TB, controlled DM with TB, and poorly-controlled DMTB patients**. Data is presented as median and interquartile range. TB n=30 TB, C_DMTB n=10 and PC_DMTB n=20. Kruskal-Wallis test with Dunn's multiple comparison test was performed. *Adjusted *p*-value<0.05, **Adjusted *p*-value<0.01, ****Adjusted *p*-value<0.0001.

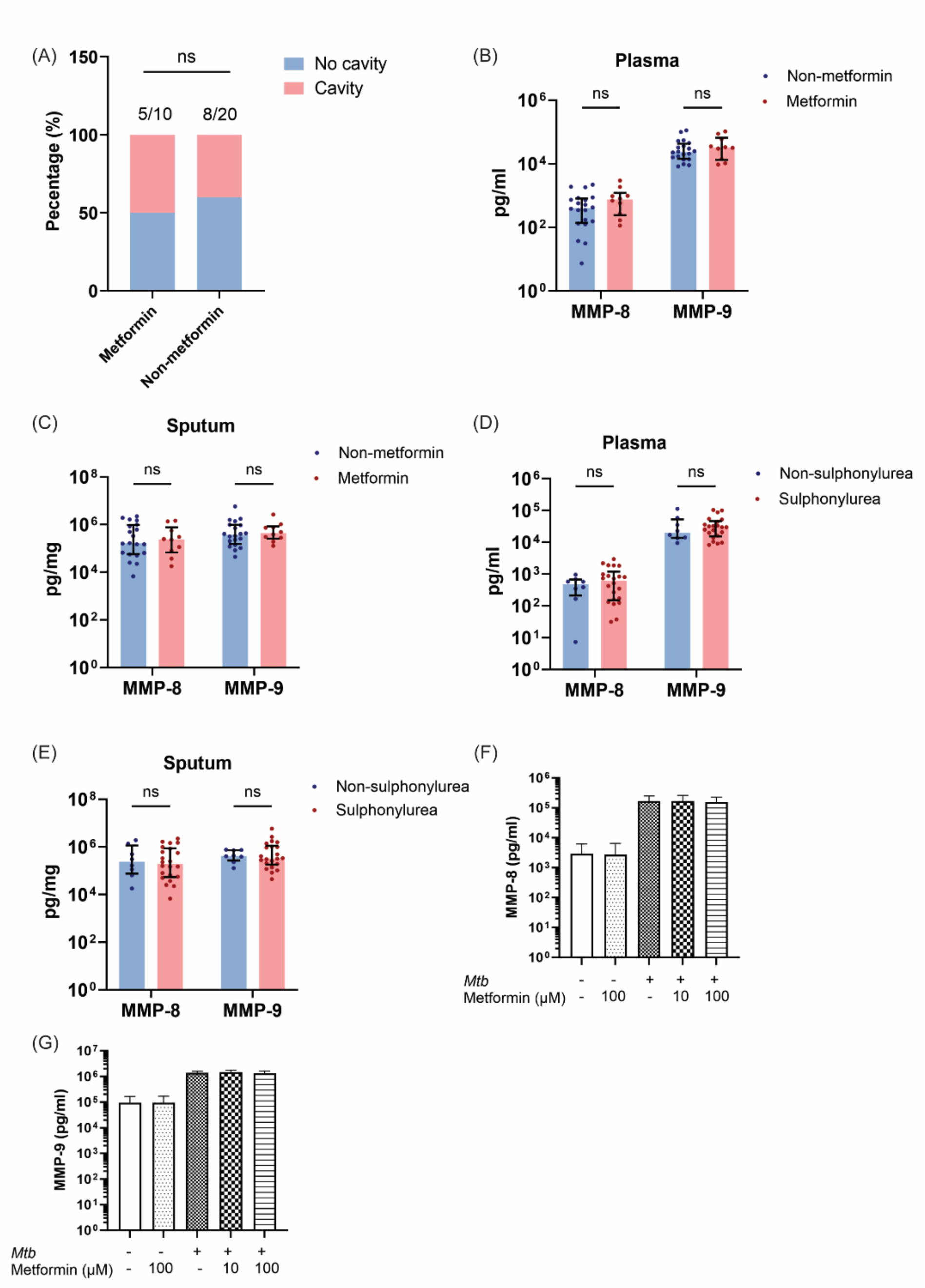

**Figure S2. Metformin and sulphonylurea treatment did not affect MMP-8 or MMP-9 in DMTB patients.** (A) Frequency of cavitation in DMTB patients on metformin and non-metformin therapy. n=10 on metformin therapy, n=20 on non-metformin therapy. Fisher's exact test was performed. (B) Plasma and (C) sputum MMP-8 and -9 concentrations in DMTB patients on metformin and non-metformin therapy. (D) Plasma and (E) sputum MMP-8 and -9 levels in DMTB patients on sulphonylurea and non-sulphonylurea therapy. n=12 sulphonylurea-treated patients, n=6 non-sulphonylurea treated patients. Multiple Mann-Whitney test was performed. Data are presented as median and interquartile range. Effects of metformin treatment on neutrophil (F) MMP-8 and (G) MMP-9 release from healthy donors in response to *Mtb* stimulation. n=3 healthy donors. One-way ANOVA test with Sidak's multiple comparison test were performed.

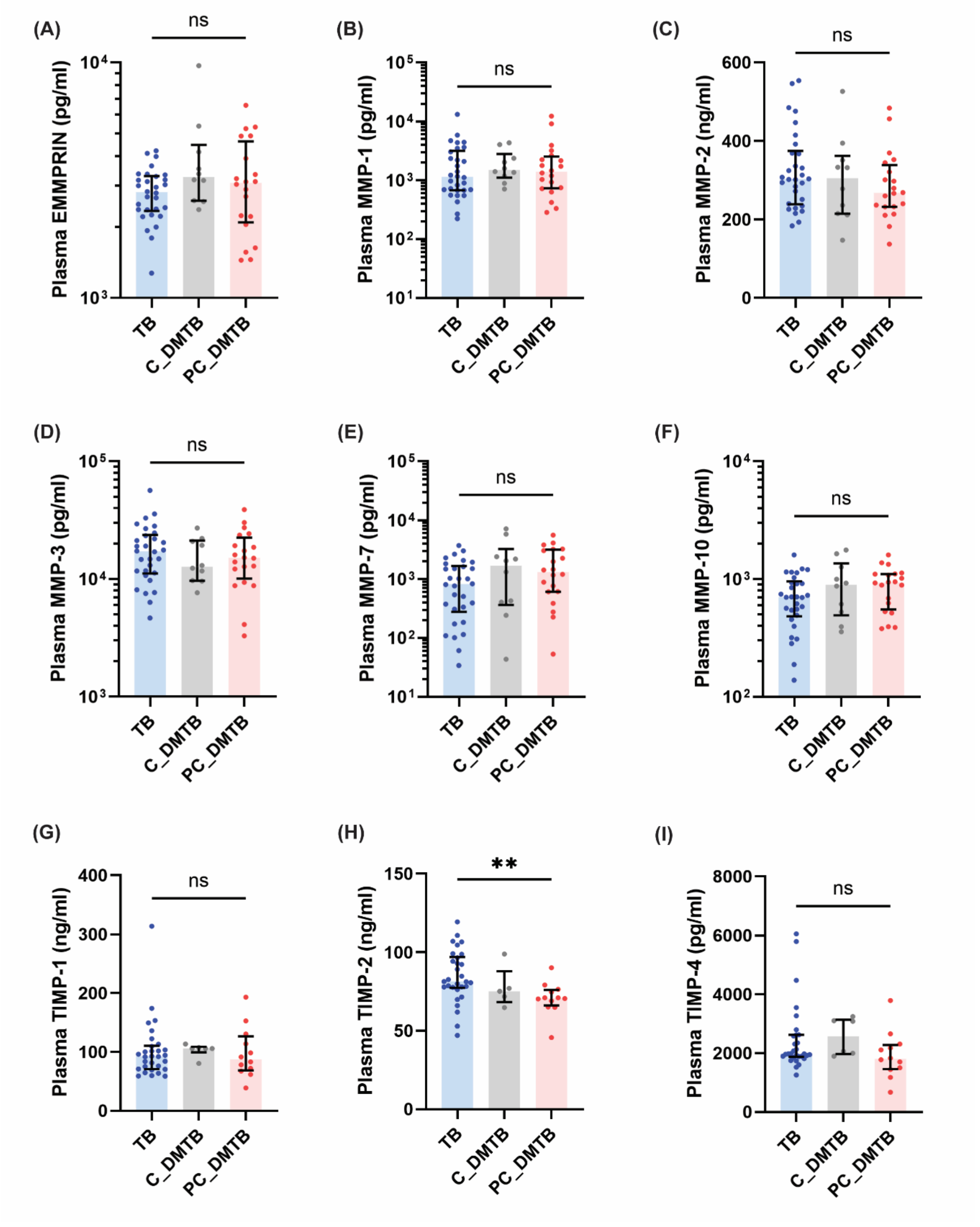

**Figure S3. Plasma EMMPRIN, MMP-1, -2, -3, -7, -10, TIMP-1, and TIMP-4 were similar across TB, C_DMTB, and PC_DMTB groups, but TIMP-2 was downregulated in PC_DMTB.** Data is presented as median and interquartile range. n=30 HC, n=30 DM, n=30 TB, n=10 C_DMTB and n=20 PC_DMTB. Kruskal-Wallis test with Dunn's multiple comparison test was performed. **Adjusted *p*-value<0.01.

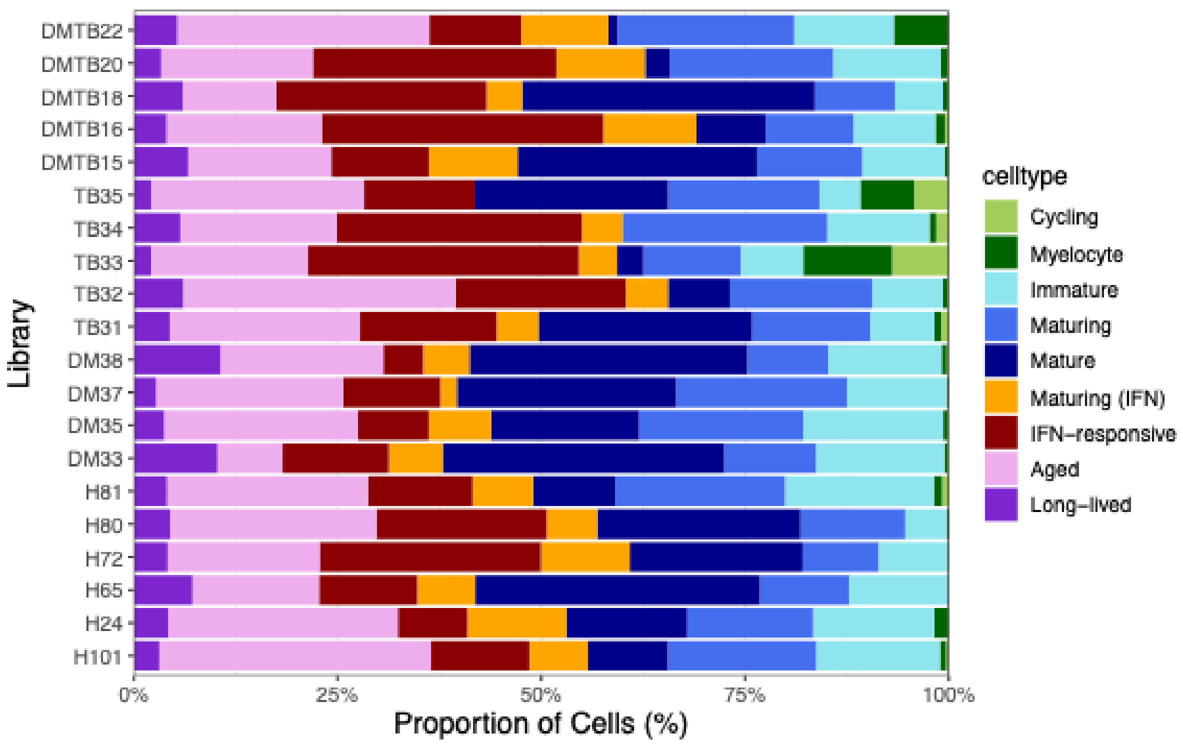

**Figure S4. Proportion of cell types for individual samples from healthy, DM, TB and DMTB groups.** Within the DMTB group, DMTB16 and DMTB22 were classified as controlled DMTB, while DMTB15, DMTB18, and DMTB20 were classified as poorly controlled DMTB. n=6 HC, n=4 DM, n=5 TB, n=5 DMTB

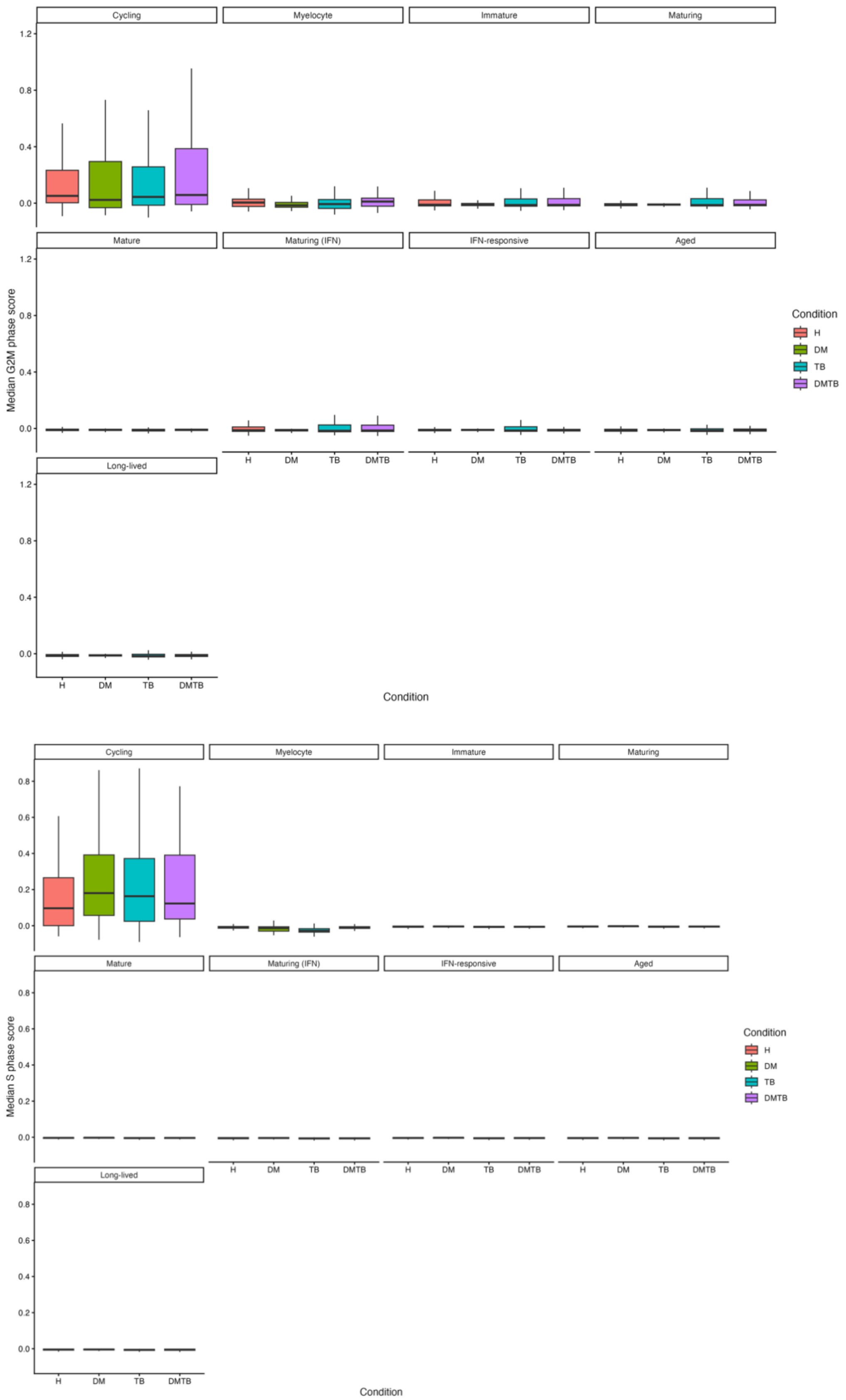

**Figure S5. Median G2 and S score using log-normalised data by cell types and conditions.**

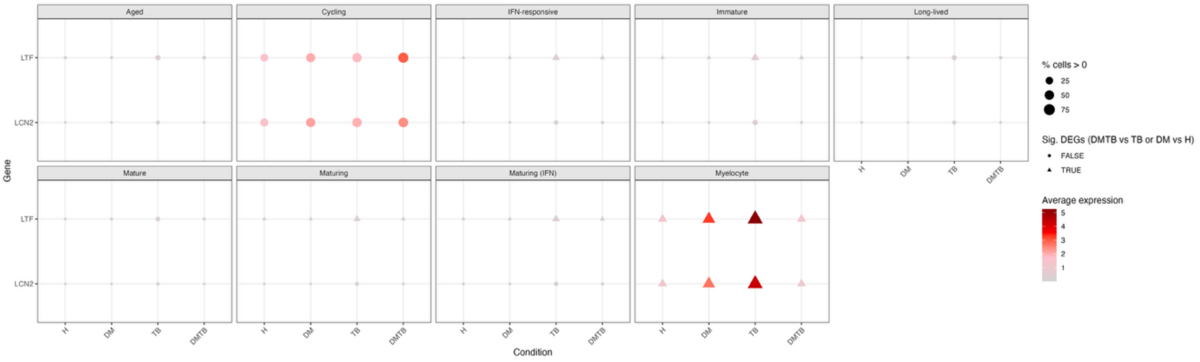

**Figure S6. Average LTF (lactoferrin) and LCN2 (lipocalin 2 or NGAL) expression using log-normalised data by cell types and conditions.** Differentially expressed genes between DMTB vs TB and DM vs H were determined using FindMarkers and using a cutoff of adjusted p-value < 0.05, min.pct > 0.1 for both conditions, FC cutoff of 1.3.

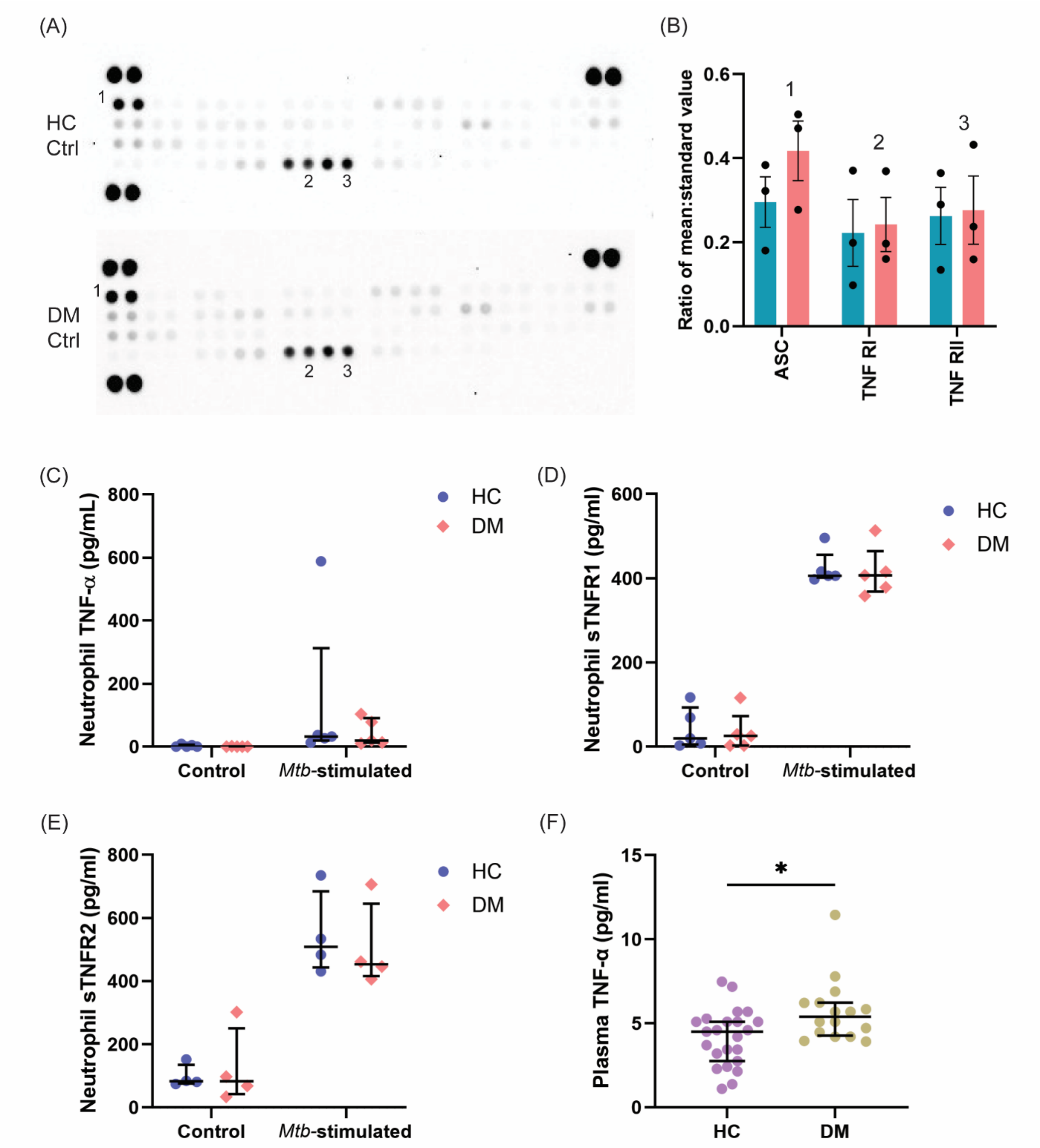

**Figure S7. Neutrophil TNF receptors and TNF-α analysis.** (A&B) NF-κB array analysis showed comparable TNFR1 and TNFR2 expression in basal unstimulated neutrophils from HC and poorly-controlled DM patients. (A) Membranes shown were representative of 3 independent donors from each group. Independent experiments yielded qualitatively identical results. (B) Densitometric analysis. Data are presented as mean ± SEM, from 3 independent donors in each group. Infection experiment was performed in pairs, consisting of 1 HC and 1 DM patient each time round. Neutrophil (C) TNF-α, (D) soluble TNFR1 and (E) soluble TNFR2 in culture supernatant 3 hours post stimulation. Two-way RM ANOVA test with Sidak multiple comparison test was performed. (F) Plasma TNF-α. n=23 HC, n=16 DM. Mann-Whitney test was performed. *$p$<0.05.

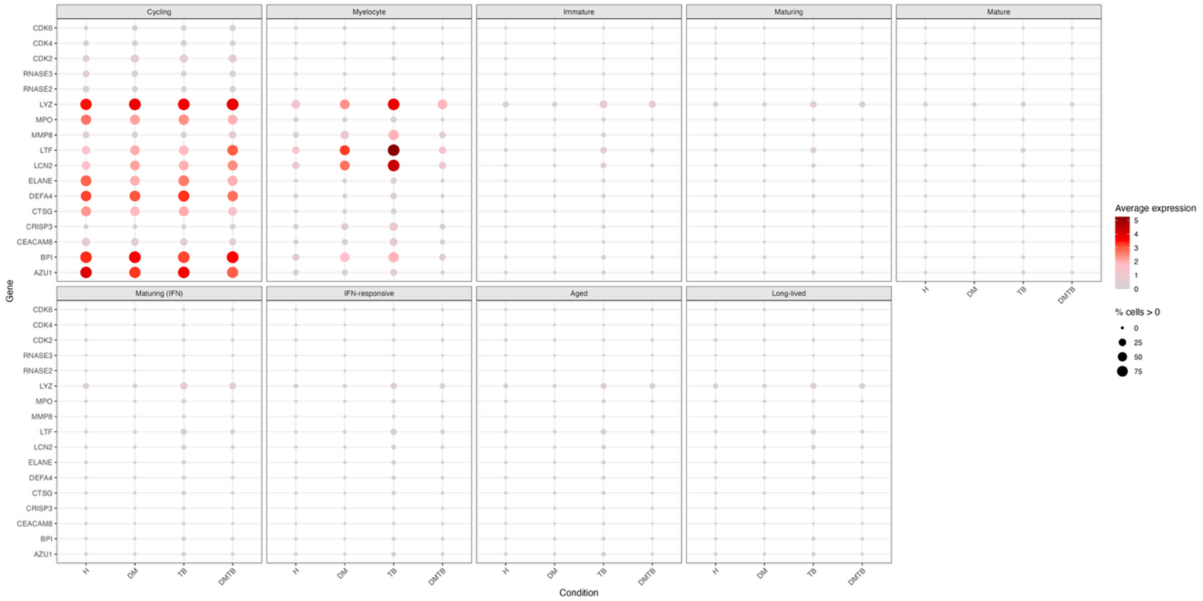

**Figure S8. Average expression of granule proteins and cell-cycle gene transcript according to (14) using log-normalised data in neutrophils.**
