## Supplementary File 1 for "Neutrophil-primed immunopathology in poorly-controlled diabetes worsens matrix destruction in pulmonary tuberculosis"

### STUDY PROTOCOL

#### Diabetes mellitus and the dysregulation of host proteases in Tuberculosis

(Version 7: 19/4/2023)

**PRINCIPAL INVESTIGATOR:**
Dr Catherine Ong, Assistant Professor, National University Health System

**STUDY SITES:**
National University Hospital, Singapore
Tan Tock Seng Hospital, Singapore

**13. Ethical Considerations……………………………………………………………………..15**
13.1. Informed Consent…………………………………………………………….......15
13.2. Confidentiality of Data and Patient Records…………………………………...15
**14. Retention of Study Documents………………………………………………………….…15**
**15. References...…………………………………………………………………………………...15**

### STUDY PROTOCOL

#### 1 BACKGROUND AND RATIONALE

Tuberculosis (TB) was declared a global health emergency in 1993 by World Health Organisation (WHO), but still remains a major global health problem today. Despite the adoption of End TB strategy by WHO, which calls for a 90% reduction in TB deaths and an 80% reduction in TB incidence rate by 2030, the annual global decline in TB incidence was only 1.5%, falling short of the 5% annual decline needed to achieve this milestone of ending TB (1). Further compounding the problem is the increasing prevalence of diabetes mellitus (DM) worldwide, including countries of high TB burden in the Asia and Western Pacific Region (7). Studies have shown that DM increases risk of active TB by threefold and is associated with higher risks of TB treatment failure and mortality (2-4). A dynamic modeling study investigating the impact of diabetes on TB in 13 high-TB burden countries estimated that if the prevalence of DM continues to rise at the present rates, we will fall well short of the End TB strategy goal (8). The convergence of both DM and TB epidemics therefore presents a major global and local public health challenge both now and in the near future, with potential to disrupt efforts to control TB in Singapore (9).

Patients with poorly controlled DM have weakened immune system, which makes them more susceptible to TB infection. Studies revealed that TB patients with DM are more likely to have higher mycobacterial load and therefore more contagious. They are also more prone to having pulmonary cavities, take a longer time to achieve sputum conversion and associated with multidrug-resistant TB (2-6). In addition, several studies demonstrated that DM-TB patients have increased subpopulations of less activated alveolar macrophages (10) and elevated peripheral blood neutrophil counts (11). Therefore, in this study, it is of our interest to investigate the effects of hyperglycaemic conditions on neutrophil and monocyte/macrophage functions and their significance in the immunopathology in DM-TB. The experimental findings obtained from this study might enable us to gain an insight into possible treatment strategies to address patient with concurrent tuberculosis and diabetes mellitus.

Tissue destruction, manifesting as lung cavitation, is a hallmark of pulmonary TB, and is the site of high mycobacterial burden. We have previously found that matrix metalloproteinase (MMP) activity is up-regulated in TB (13). Over activity of these proteases relative to their specific tissue inhibitors, tissue inhibitor of metalloproteinases (TIMPs) (12), resulting in a matrix degradation phenotype. A study conducted by Nakamura and his peers revealed that 71% of patients with concurrent tuberculosis and diabetes mellitus have cavities as compared to 45.5% of TB patients without DM (14), which strongly implicates dysregulated MMP concentrations or functional activity in DM. Human serine proteases, another component that drives matrix destruction, are also upregulated in TB and their activity contribute to the control of *Mtb* in granulomas (15). In this study, secretion of MMPs and serine proteases in DM-TB patients will be evaluated. We will also analyse measures of resultant tissue destruction and immunopathology, such as matrix degradation products and inflammatory mediators associated with lung pathology.

In summary, the current convergence of both the DM and TB epidemics globally highlights the urgent need to address the clinical issue of DM-TB. The underlying mechanisms causing a more severe disease phenotype in DM-TB are not clearly defined. Investigating the mechanisms driving DM-associated TB immunopathology in this study will provide new knowledge that can improve management of DM-TB patients, and may lead to host-directed therapy that can optimise patient management and potentially contribute significantly towards ending TB.

### 2 HYPOTHESIS AND OBJECTIVES

#### 2.1 Hypothesis

Diabetes mellitus (DM) modulates host proteases driving TB immunopathology.

#### 2.2 Aims

These are to define the immunopathology of TB in DM by investigating:

a) the gene expression, secretion and functional activity of matrix metalloproteinases (MMPs) and serine proteases in hyperglycemic conditions
b) the gene expression, secretion and functional activity of MMPs and serine proteases in TB patients with and without DM
c) the intracellular signalling pathways regulating MMPs and serine proteases in hyperglycemic conditions

### 3 STUDY MANAGEMENT

We will appoint co-ordinators to supervise the day-to-day running of the study, compliance, recruitment, budget control and problem solution. Study management shall be coordinated by the Investigational Medicine Unit at the National University Hospital and Singapore Infectious Diseases Clinical Research Network (SCRN).

### 4 STUDY SCHEDULE

**DM patients**[6]

| Visit timing[1] | Scr | D0 |
|---|---|---|
| Entry criteria | X | |
| Informed consent | X | |
| Medical history | X | |
| Urine Pregnancy test[2] | X | |
| Blood for HbA1c test[3] | X | |
| Blood collection[4] | | X |
| Urine collection[5] | | X |

1. D0 visit to be done as soon as possible after screening (can be the same day).
2. Urine pregnancy test only needed for women of childbearing potential.
3. HbA1c test need not be repeated if done in previous 90 days.
4. Blood will be analysed for biological factors and immune profiles. Neutrophils and monocytes isolated will be stimulated and analysed.
5. Urine will be analysed for biological factors, immune profiles and archived for future mass spectrometry.
6. DM patients may be required to fast a minimum of 6 hours prior to blood collection. DM patients will be invited back to the study, no often than once every fortnight. The volume of blood drawn will not exceed 180ml in a 90-day period. HbA1c screening test will be repeated if the result is no longer valid.

TB patients[9]

| Visit timing[1] | Scr | D0 |
|---|---|---|
| Entry criteria | X | |
| Informed consent | X | |
| Standard combination TB treatment | (X) | X |
| Medical history | X | |
| Physical examination (targeted, for new symptoms) | X | |
| Urine Pregnancy test[2] | X | |
| CXR[3] | X | |
| Sputum for GeneXpert test[4] | X | |
| Blood for HIV and HbA1c[5] | X | |
| Blood collection[6] | | X |
| Urine collection[7] | | X |
| Induced sputum collection[8] | | X |

1. D0 visit to be done as soon as possible after screening (can be same day).
2. Urine pregnancy test only needed for women of childbearing potential.
3. CXR is not required if done in previous 14 days and results available to the study team.
4. GeneXpert test need not be repeated if done in previous 14 days and results available to the study team. GeneXpert not required if sputum is smear positive and CXR is in keeping with PTB or sputum TB cultures grew *M. tuberculosis.*
5. HbA1c need not be repeated if done in previous 90 days. HIV antibody test need not be repeated if done in previous 90 days or the result is known to be positive.
6. Blood will be analysed for biological factors and immune profiles.
7. Urine will be analysed for biological factors, immune profiles and archived for future mass spectrometry.
8. Induced sputum samples will be analysed for biological factors and immune profiles.
9. TB patients may be required to fast a minimum of 6 hours prior to blood collection.

**DM-TB patients[9]**

| Visit timing[1] | Scr | D0 |
|---|---|---|
| Entry criteria | X | |
| Informed consent | X | |
| Standard combination TB treatment | (X) | X |
| Medical history | X | |
| Physical examination (targeted, for new symptoms) | X | |
| Urine Pregnancy test[2] | X | |
| CXR[3] | X | |
| Sputum for GeneXpert test[4] | X | |
| Blood for HIV and HbA1c[5] | X | |
| Blood collection[6] | | X |
| Urine collection[7] | | X |
| Induced Sputum collection[8] | | X |

1. D0 visit to be done as soon as possible after screening (can be same day).
2. Urine pregnancy test only needed for women of childbearing potential.
3. CXR is not required if done in previous 14 days and result is available to the study team.

4. GeneXpert test need not be repeated if done in previous 14 days and result is available to the study team. GeneXpert not required if sputum is smear positive and CXR is in keeping with PTB or sputum TB cultures grew M. tuberculosis.
5. HbA1c need not be repeated if done in previous 90 days. HIV antibody test need not be repeated if done in previous 90 days or the result is known to be positive.
6. Blood will be analysed for biological factors and immune profiles.
7. Urine will be analysed for biological factors, immune profiles and archived for future mass spectrometry.
8. Induced sputum samples will be analysed for biological factors and immune profiles.
9. DM-TB patients may be required to fast a minimum of 6 hours prior to blood collection.

### 5 STUDY POPULATION

#### 5.1 Selection of Patients

##### 5.1.1 DM patients

52 type 2 diabetic patients will be recruited from NUH, with no gender restriction.

***Inclusion criteria:***

1. Aged 21 years to less than 70.
2. Uncontrolled type II DM with HbA1c > 8%

***Exclusion criteria:***

1. Unable to give informed consent
2. Prisoners
3. Pregnancy
4. Chronic kidney disease - Creatinine 2 times upper limit of normal (ULN)
5. Autoimmune disease on systemic immunosuppressant
6. Any concurrent acute illness, such as influenza
7. Principal investigator assessment of lack of willingness to participate and comply with all requirements of the protocol, or identification of any factor felt to significantly increase the participant's risk of suffering an adverse outcome

The same 52 DM patients will be invited back to the study, no often than once every fortnight, if their HbA1c results satisfy the eligibility criteria and are within the validity period. Each HbA1c test result is valid for 90 days. Multiple visits is required for DM patients due to the different downstream assessment between DM, TB and DM-TB patients. One of the aims of this study is to investigate the effects of hyperglycaemic condition on immune cell function and their significance in the immunopathology of DM-TB. To achieve this aim, immune cells will be isolated from the blood of DM patients, stimulated with live M. bovis BCG or live virulent M. tuberculosis H37Rv and assessed for their function. A larger volume of blood is required to complete these assessments, which explains why DM patients are requested to make multiple visits.

While multiple visits is required for DM patients, the investigators will assess the participants' health status to determine if they are suitable for the blood draw during each visit. If there is any factor felt to significantly increase participant's risk of suffering an adverse outcome, he or she will be excluded from the blood draw. The study team will ensure that the frequency of visit does not exceed the volume of blood drawn over a stated period, which is 180 mL within a 90-

day period.

##### 5.1.2 TB patients

TB patients will be recruited from National University Hospital (NUH), Tan Tock Seng Hospital and Luyang TB Clinic, Sabah, Malaysia with no gender restriction. The clinical site must be involved in the treatment of patients with tuberculosis. The investigator must have appropriate experience of conducting trials according to Good Clinical Practice. The site must have adequate staff and facilities to be able to conduct the study properly.

***Inclusion criteria:***

1. Aged 21 years to 99 years
2. Patients receiving ≤ 7 days of TB treatment or about to start standard combination TB treatment
3. Confirmed pulmonary TB with positive acid-fast bacilli smear and/or positive GeneXpert and/or culture results
4. CXR demonstrating pulmonary involvement

***Exclusion criteria:***

1. Diabetes mellitus
2. Unable to give informed consent
3. Pregnancy
4. Prisoners
5. HIV co-infection
6. Previous pulmonary TB
7. Severe, pre-existing lung diseases
8. Autoimmune disease and/or on systemic immunosuppressant
9. Chronic kidney disease - Creatinine 2 times upper limit of normal (ULN)
10. Principal investigator assessment of lack of willingness to participate and comply with all requirements of the protocol, or identification of any factor felt to significantly increase the participant's risk of suffering an adverse outcome

##### 5.1.3 DM-TB patients

DM-TB patients will be recruited from National University Hospital (NUH), Tan Tock Seng Hospital and Luyang TB Clinic, Sabah, Malaysia with no gender restriction. The clinical site must be involved in the treatment of patients with tuberculosis. The investigator must have appropriate experience of conducting trials according to Good Clinical Practice. The site must have adequate staff and facilities to be able to conduct the study properly.

***Inclusion criteria:***

1. Aged 21 years to 99 years
2. Patients receiving ≤ 7 days of TB treatment or about to start standard combination TB treatment
3. Confirmed pulmonary TB with positive acid-fast bacilli smear and/or positive GeneXpert and/or culture results
4. CXR demonstrating pulmonary involvement
5. Type II DM with HbA1c ≥6.5%

***Exclusion criteria:***

1. Unable to give informed consent
2. Pregnancy
3. Prisoners
4. HIV co-infection
5. Previous pulmonary TB
6. Severe, pre-existing lung diseases
7. Autoimmune disease and/or on systemic immunosuppressant
8. Chronic kidney disease - Creatinine 2 times upper limit of normal (ULN)
9. Principal investigator assessment of lack of willingness to participate and comply with all requirements of the protocol, or identification of any factor felt to significantly increase the participant's risk of suffering an adverse outcome

It is noted that the number of TB and DM-TB patients recruited are not fixed for each site, but will make up to a total of 30 patients for each group for recruitment in Singapore (both NUH and Tan Tock Seng Hospital). For recruitment in Sabah Malaysia, additional 30 TB and 30 DM-TB patients will be recruited.

**Note:**
The recruitment for TB patients without DM will be increased to 35. As of 19 April 2023, all 30 TB patients have been recruited. However, the study team would like to conduct single cell analysis on immune cells isolated from TB patients. Therefore would like to recruit additional 5 patients.

#### 5.2 Withdrawal from the Study and Subject Replacement

DM, TB and DM-TB patients could be withdrawn from the study for any of the following reason:

1. Patient withdrawal of consent to participate in the study
2. Patient no longer satisfies the eligibility criteria (eg. DM patient's HbA1c level falls below 8%)
3. Principal investigator or managing physicians considers it to be in the best interests of the patient to withdraw from the study.

Any participant who withdraws from the study will be replaced.

### 6 SCREENING

All potential participants will be screened. Prior to taking consent for screening or performing any screening related procedures, a check of medical and drug history will be performed to ensure that the patient meets basic clinical eligibility criteria. In NUH and TTSH, investigators will access the individual's electronic medical records for the initial pre-screening and screening. Participants who appear suitable on this initial pre-screening will be given adequate information about the study together with a participant information sheet and given an opportunity to ask questions about the study. If the patient is willing to proceed, then they must sign the consent form. A thumbprint is acceptable for patients who are unable to provide a signature.

#### 6.1 DM patients

Screening assessments will be performed as listed below and as summarised in the study schedule:

- Review of medical history
- Urine pregnancy test for women of child-bearing potential
- Blood test: HbA1c (HbA1c test need not be repeated if done in previous 90 days)

#### 6.2 TB patients

Screening assessments will be performed as listed below and as summarised in the study schedule:

- Review of medical history
- Review of current symptoms
- Physical examination (vital signs and targeted examination to evaluate reported symptoms)
- Chest X-ray (need not be repeated if performed within the previous 14 days and the film is available for evaluation by the research team).
- Sputum collection for GeneXpert test to confirm *M. tuberculosis* and for rifampicin sensitivity (need not be repeated if done within the previous 14 days and results available to study team). GeneXpert is not required if AFB smear is positive and CXR is in keeping with pulmonary TB or sputum TB cultures grew *M. tuberculosis*
- Urine pregnancy test for women of child-bearing potential
- Blood tests: HbA1c and HIV (HIV tests need not be repeated if done in previous 90 days or when HIV test is known to be positive; HbA1c need not be repeated if done in previous 90 days)

#### 6.3 DM-TB patients

Screening assessments will be performed as listed below and as summarised in the study schedule:

- Review of medical history
- Review of current symptoms
- Physical examination (vital signs and targeted examination to evaluate reported symptoms)
- Chest X-ray (need not be repeated if performed within the previous 14 days and the film is available for evaluation by the research team).
- Sputum collection for GeneXpert test to confirm *M. tuberculosis* and for rifampicin sensitivity (need not be repeated if done within the previous 14 days and results available to study team). GeneXpert is not required if AFB smear is positive and CXR is in keeping with pulmonary TB or sputum TB cultures grew *M. tuberculosis.*
- Urine pregnancy test for women of child-bearing potential
- Blood tests: HbA1c and HIV (HIV tests need not be repeated if done in previous 90 days or when HIV test is known to be positive; HbA1c need not be repeated if done in previous 90 days)

### 7 SAMPLE AND DATA COLLECTION

Samples are collected at Day 0 visit. Day 0 visit can take place on the same day of the screening visit. For TB and DM-TB patients, Day 0 may be deferred to another day as long as the patient is within 7 days of initiating TB treatment.

#### 7.1 Sample collection

##### 7.1.1 DM patients

The following procedures will be performed at the Day 0 visit:

- Up to 60mls of blood and 20mls of urine will be collected

DM patients will be invited back to the study, no often than once every fortnight, as long as they satisfy the eligibility criteria. The volume of blood drawn will not exceed 180ml in a 90-day period. The following procedures will be performed at subsequent visit:

- Up to 60mls of blood will be collected. Urine may be collected, dependent on the Investigator's discretion

##### 7.1.2 TB and DM-TB patients

The following procedures will be performed at the Day 0 visit:

- Up to 60mls of blood, 20mls of urine and 10mls of induced sputum will be collected

**Induced sputum collection for TB and DM-TB patients**

Sputum induction will be performed in accordance with a locally agreed standard operating procedure which is already in use for routine clinical care. All induction will be performed in a designated collection room according to protocols and will be supervised by a member of the study team to ensure participant safety, consistency and adequacy of sample collection and ensure no cross-contamination occurs.

The patient will be unaccompanied in the room during the 5 minute periods of nebulisation, but kept under observation. At the end of nebulisation, the subject will remain in the room until coughing has ceased, after which they will be free to leave and the room will be decontaminated. Subjects who are considered to be infectious will be requested to wear a respiratory protective mask until they have either left the hospital building or returned to an isolation room on the ward. Researchers will wear N95 masks whilst assisting the patient with the sputum induction procedure. We have utilised this induced sputum procedure for other research studies with no significant adverse events (16, 17).

#### 7.2 Data collection

Investigators will access NUH and TTSH participants' electronic medical records for collection of clinical information. All collected clinical information will be identified only by the research number and patient initials. Clinical data such as time to sputum sterilisation, type of ATTs, duration of treatment, type of oral hypoglycaemic agents the patient is on (metformin, thiazolidinedione or others), date or year of diagnosis of DM will be recorded. Data collected in the case report form can also be used as clinical information.

### 8 SAMPLE ANALYSIS

#### 8.1 DM patients

##### 8.1.1 Whole blood

Plasma will be processed from the whole blood, and analysed for biological and immune factors such as cytokines, chemokines, circulating *M. tuberculosis* DNA, proteases and matrix degradation products. Immune cells such as neutrophils and monocytes will be extracted. Monocytes will be matured to macrophages. Immune cells will be assessed for their immune functions and signalling pathways in response to mycobacteria infection. The culture supernatants will be analysed for biological and immune factors. Other blood components including platelets will be extracted and analysed. Whole blood will also be sent for transcriptomics analysis. Single cell RNA sequencing will also be performed. Full blood count test might be performed to determine leukocytes count in the blood.

##### 8.1.2 Urine

Urine samples collected will be analysed for biological factors such as matrix degradation products and circulating *Mtb* DNA, and will also be archived for future mass spectrometry.

#### 8.2 TB patients and DM-TB patients

##### 8.2.1 Whole blood

Plasma will be processed from the whole blood in a Biosafety Level 3 (BSL3) containment facility and will be inactivated in accordance to BSL3 inactivation protocols, before transferring out to BSL2 laboratory. Inactivated plasma will be analysed for biological and immune factors such as matrix metalloproteinases (MMPs), cytokines, chemokines and circulating *Mtb* DNA. Whole blood will also be sent for transcriptomic analysis. Immune cells including leukocytes and platelets will be extracted and infected with mycobacteria. The immune cells will be assessed for their biological functions. Full blood count test might be performed to determine leukocytes count in the blood.

##### 8.2.2 Urine

Urine will be inactivated in the BSL3 facility in accordance to BSL3 inactivation protocols, before transferring out to BSL2 laboratory. Inactivated urine will be analysed for biological factors and will also be archived for future mass spectrometry.

##### 8.2.3 Sputum

Clinical samples from the site of infection (eg. sputum, lymph node, etc) are collected from TB patients and cultured in TB lab as part of the routine clinical care for TB infection screening. *Mtb* isolates will be retrieved from NUH laboratory for downstream analysis.

Sputum samples obtained will be inactivated in accordance to BSL3 inactivation protocols, before transferring out to BSL2 laboratory. Inactivated sputum will be analysed for biological and immune factors.

### 9 MEDICAL MANAGEMENT

**TB patients and DM-TB patients** will be managed as an outpatient during the study, although in selected cases patients will be managed as inpatients according to the primary medical team. Patients managed in the outpatient setting will be given an emergency contact number of the study team and will be given clear instructions on how to access medical care from appropriate facilities in the event of a medical emergency.

Complications of TB will be managed as clinically appropriate. Other medical conditions will be managed by the site study team as clinically appropriate.

**Diabetic patients** will be managed as outpatients during the period of the study.

### 10 SOCIAL/ECONOMIC AND ETHICAL IMPLICATIONS

#### 10.1 Financial implications

Participants will be compensated $50 per visit for their inconvenience, discomfort and travel expenses when contributing to the study. Tests performed as part of screening will be costed to the study. There will be no additional financial cost for participants in addition to that incurred by their standard outpatient/hospital care.

#### 10.2 Time expected for participation

Obtaining clinical samples (blood, urine and induced sputum) is estimated to take less than 30 minutes. There will be adequate, indefinite time allowed in all cases for patients to consider informed consent. The required tests should all take place during one attendance at the medical facility, removing the need for patients to return to the clinical site on a second occasion. Participation will not otherwise interfere with routine clinical care.

#### 10.3 Benefits to participants/patients

It is not envisaged that participants will benefit overtly from participating in the study, however the result of the study may influence future TB management and indirectly benefit the patient population by improving clinical care.

#### 10.4 Potential adverse effects of researchers

1) Working with clinical blood samples: Researchers are experienced in handling blood samples and have been vaccinated against hepatitis B with detectable antibody levels. Routine universal precautions (wearing gloves, no use of sharps after phlebotomy) will be employed to minimise the risk of HIV transmission.
2) Sputum induction: Due to aerosolising of respiratory secretions there is a theoretical risk of increased transmission of TB. However this is minimised by physical isolation in the induced sputum facility during the sputum induction procedure and use of N95 masks by the patients and researchers at appropriate times.

#### 10.5 Responsibilities of the ethics committee and investigator

Before starting this study, the following requirements must be satisfied. The protocol and the informed consent document must be submitted, reviewed and approved by an Independent Ethics Committee responsible for each study site. A signed and dated statement that the protocol and informed consent have been approved by the Ethics committee must be given to Principal Investigator, IMU, SCRN and NUH RO before study initiation.

The investigator must sign a protocol signature page confirming his/her agreement to conduct the study in accordance with these documents (study instructions and procedures found in this protocol), and to provide access to all relevant data and records to field monitors, Quality Assurance representatives and designees from IMU, SCRN, auditors, ethics committees and regulatory authorities as required. In the event there is an inspection of the clinical site requested by a regulatory authority, the investigator must inform IMU, SCRN and NUH RO immediately that this request has been made.

Investigators must adhere to the protocol and its procedures to avoid protocol deviations. The investigator shall not contact Principal Investigator to request approval of a protocol deviation because there are no allowable authorized deviations. If the investigator feels a protocol deviation would improve the conduct of the study, this must be considered as a protocol amendment. Such an amendment must be agreed upon by Principal Investigator and shall be approved by the Ethics Committee before implementation.

Any change or addition to the protocol can only be made in a written protocol amendment that must be approved by Principal Investigator, Health Authorities where required, and the Ethics committee. Only amendments that are required for patient safety may be implemented prior to ethics committee approval. Notwithstanding the need for approval of formal protocol amendment, the investigator is expected to take any immediate action required for the safety of any patient included in this study, even if this action represents a deviation from the protocol. In such cases, the Principal Investigator, IMU, SCRN and NUH RO should be notified of this action and the Ethics committee at the study site should be informed in accordance to the requirements stated in ethics committee standard operating procedures, if applicable, to be reported as per local regulatory requirements.

### 11 DATA ENTRY AND STORAGE

Research data will be stored inside excel spread sheet with password protection and restricted access. The files will be encrypted. The laptop will be locked and kept in a facility in which users have card access. Patients' information, including their medical record and drug history will be maintained by IMU and Singapore Clinical Research Network.

### 12 SAMPLE SIZE AND STATISTICAL METHODS

The primary objective of the study is to compare the five analytes (MMP-3, -10, EMPRIN, PIIINP and desmosine) between TB patients with and without DM. 60 subjects (30 TB patients with diabetes and 30 TB patients without diabetes) will be recruited. Assuming the mean difference of the selected variables between 2 groups of patients is about 1 standard deviation, the sample size is sufficient to detect a difference with 80% power and the significance level was chosen at 0.01. Data will be analysed using Graphpad Prism software, using Mann-Whitney U test for non-parametric variables and two-way ANOVA for comparisons across the DM-TB and non-DM TB patient followed by Bonferroni test.

### 13 ETHICAL CONSIDERATIONS

#### 13.1 Informed Consent

Recruitment of participants would be done after they have undergone screening procedures, consisting of medical history check and medical test, read the information sheet and signed the consent form. A thumbprint is acceptable for patients who are unable to provide a signature. For non-English speaking participants, a translator that is fluent in both English and the participants' spoken language will be present to explain the study to them and answer their queries.

#### 13.2 Confidentiality of Data and Patient Records

Patients' identification log will be kept securely by IMU and SCRN. To maintain participants' confidentiality, all samples collected will be coded before sending to NUS laboratory for analysis. Samples will be identified only by the research number and participants' initials. In the event of any publication regarding this study, patient's identity will remain confidential.

### 14 RETENTION OF STUDY DOCUMENTS

Documents will be stored in accordance of Good Clinical Practice Guidelines (GCP) guidelines for about 6 years after the completion of the study.
