## Supplementary File 2 for "Neutrophil-primed immunopathology in poorly-controlled diabetes worsens matrix destruction in pulmonary tuberculosis"

| ID | Description | GeneRatio | BgRatio | RichFactor | ldEnrichme | zScore | pvalue | p.adjust | qvalue | geneID | Count |
|---|---|---|---|---|---|---|---|---|---|---|---|
| GO:00096 | response to bacterium | 20/72 | 451/11391 | 0.044346 | 7.015891 | 10.39689 | 2.74E-12 | 5.2E-09 | 4.6E-09 | FOS/LTF/L | 20 |
| GO:00427 | defense response to bacterium | 11/72 | 180/11391 | 0.061111 | 9.668287 | 9.349135 | 1.48E-08 | 1.4E-05 | 1.24E-05 | LTF/LCN2/ | 11 |
| GO:00019 | cell killing | 10/72 | 154/11391 | 0.064935 | 10.27327 | 9.240428 | 3.98E-08 | 2.52E-05 | 2.23E-05 | LTF/DEFA3 | 10 |
| GO:00096 | response to virus | 13/72 | 332/11391 | 0.039157 | 6.194905 | 7.661482 | 1.3E-07 | 6.18E-05 | 5.47E-05 | DEFA3/IFI4 | 13 |
| GO:00316 | killing of cells of another organism | 6/72 | 48/11391 | 0.125 | 19.77604 | 10.39644 | 5.13E-07 | 0.00014 | 0.000124 | LTF/DEFA3 | 6 |
| GO:01410 | disruption of cell in another organism | 6/72 | 48/11391 | 0.125 | 19.77604 | 10.39644 | 5.13E-07 | 0.00014 | 0.000124 | LTF/DEFA3 | 6 |
| GO:00516 | defense response to virus | 11/72 | 255/11391 | 0.043137 | 6.824673 | 7.502416 | 5.16E-07 | 0.00014 | 0.000124 | DEFA3/IFI4 | 11 |
| GO:01500 | regulation of neuroinflammatory response | 5/72 | 28/11391 | 0.178571 | 28.25149 | 11.51455 | 7.69E-07 | 0.000176 | 0.000156 | PTGS2/PTP | 5 |
| GO:01410 | disruption of anatomical structure in anoth | 6/72 | 52/11391 | 0.115385 | 18.25481 | 9.946001 | 8.35E-07 | 0.000176 | 0.000156 | LTF/DEFA3 | 6 |
| GO:00712 | cellular response to molecule of bacterial o | 9/72 | 174/11391 | 0.051724 | 8.18319 | 7.615136 | 1.37E-06 | 0.00026 | 0.00023 | LTF/DEFA3 | 9 |
| GO:00485 | negative regulation of viral process | 6/72 | 64/11391 | 0.09375 | 14.83203 | 8.849986 | 2.9E-06 | 0.00045 | 0.000398 | LTF/APOB | 6 |
| GO:00326 | tumor necrosis factor production | 8/72 | 143/11391 | 0.055944 | 8.850816 | 7.534757 | 3.08E-06 | 0.00045 | 0.000398 | LTF/CYBB/ | 8 |
| GO:00326 | regulation of tumor necrosis factor product | 8/72 | 143/11391 | 0.055944 | 8.850816 | 7.534757 | 3.08E-06 | 0.00045 | 0.000398 | LTF/CYBB/ | 8 |
| GO:00717 | tumor necrosis factor superfamily cytokine | 8/72 | 147/11391 | 0.054422 | 8.609977 | 7.406375 | 3.79E-06 | 0.000479 | 0.000424 | LTF/CYBB/ | 8 |
| GO:19035 | regulation of tumor necrosis factor superfa | 8/72 | 147/11391 | 0.054422 | 8.609977 | 7.406375 | 3.79E-06 | 0.000479 | 0.000424 | LTF/CYBB/ | 8 |
| GO:00712 | cellular response to biotic stimulus | 9/72 | 200/11391 | 0.045 | 7.119375 | 6.963245 | 4.34E-06 | 0.000514 | 0.000455 | LTF/DEFA3 | 9 |
| GO:00450 | regulation of viral genome replication | 6/72 | 71/11391 | 0.084507 | 13.36972 | 8.338537 | 5.34E-06 | 0.000596 | 0.000527 | LTF/APOB | 6 |
| GO:00022 | response to molecule of bacterial origin | 10/72 | 268/11391 | 0.037313 | 5.903296 | 6.478413 | 6.52E-06 | 0.000687 | 0.000608 | FOS/LTF/D | 10 |
| GO:00450 | negative regulation of viral genome replica | 5/72 | 45/11391 | 0.111111 | 17.5787 | 8.887089 | 8.8E-06 | 0.000817 | 0.000722 | LTF/APOB | 5 |
| GO:00712 | cellular response to lipopolysaccharide | 8/72 | 165/11391 | 0.048485 | 7.670707 | 6.883755 | 8.91E-06 | 0.000817 | 0.000722 | LTF/DEFA3 | 8 |
| GO:00507 | regulation of viral process | 7/72 | 118/11391 | 0.059322 | 9.38524 | 7.30232 | 9.04E-06 | 0.000817 | 0.000722 | LTF/APOB | 7 |
| GO:01403 | antiviral innate immune response | 5/72 | 51/11391 | 0.098039 | 15.51062 | 8.283019 | 1.64E-05 | 0.001416 | 0.001253 | EIF2AK2/O | 5 |
| GO:00327 | positive regulation of tumor necrosis factor | 6/72 | 92/11391 | 0.065217 | 10.31793 | 7.15678 | 2.4E-05 | 0.001977 | 0.001749 | CYBB/PTP | 6 |
| GO:00069 | humoral immune response | 7/72 | 138/11391 | 0.050725 | 8.02506 | 6.621853 | 2.51E-05 | 0.001987 | 0.001757 | LTF/DEFA3 | 7 |
| GO:00324 | response to lipopolysaccharide | 9/72 | 252/11391 | 0.035714 | 5.650298 | 5.953632 | 2.78E-05 | 0.002011 | 0.001779 | FOS/LTF/D | 9 |
| GO:19035 | positive regulation of tumor necrosis factor | 6/72 | 95/11391 | 0.063158 | 9.992105 | 7.019156 | 2.88E-05 | 0.002011 | 0.001779 | CYBB/PTP | 6 |
| GO:19039 | regulation of viral life cycle | 6/72 | 95/11391 | 0.063158 | 9.992105 | 7.019156 | 2.88E-05 | 0.002011 | 0.001779 | LTF/APOB | 6 |
| GO:00018 | positive regulation of cytokine production | 11/72 | 389/11391 | 0.028278 | 4.473757 | 5.559843 | 2.97E-05 | 0.002011 | 0.001779 | CHI3L1/PT | 11 |
| GO:00190 | viral genome replication | 6/72 | 107/11391 | 0.056075 | 8.871495 | 6.524416 | 5.64E-05 | 0.003671 | 0.003247 | LTF/APOB | 6 |
| GO:01500 | neuroinflammatory response | 5/72 | 66/11391 | 0.075758 | 11.98548 | 7.138319 | 5.81E-05 | 0.003671 | 0.003247 | PTGS2/PTP | 5 |
| GO:00197 | antimicrobial humoral response | 5/72 | 67/11391 | 0.074627 | 11.80659 | 7.075389 | 6.24E-05 | 0.00382 | 0.003379 | LTF/DEFA3 | 5 |
| GO:00507 | negative regulation of immune response | 7/72 | 162/11391 | 0.04321 | 6.836163 | 5.966758 | 7.01E-05 | 0.004155 | 0.003675 | PGLYRP1/P | 7 |
| GO:00313 | negative regulation of defense response | 7/72 | 209/11391 | 0.033493 | 5.298844 | 5.002523 | 0.00034 | 0.016954 | 0.014996 | PGLYRP1/P | 7 |
| GO:00319 | response to corticosteroid | 5/72 | 97/11391 | 0.051546 | 8.155069 | 5.644167 | 0.00036 | 0.0175 | 0.015479 | FOS/PTGS | 5 |
| GO:00321 | negative regulation of response to external | 8/72 | 291/11391 | 0.027491 | 4.34937 | 4.616068 | 0.000478 | 0.022095 | 0.019543 | LTF/PGLYR | 8 |
| GO:00028 | negative regulation of response to biotic sti | 5/72 | 104/11391 | 0.048077 | 7.60617 | 5.397607 | 0.000496 | 0.02241 | 0.019821 | LTF/USP15 | 5 |
| GO:00457 | positive regulation of angiogenesis | 5/72 | 110/11391 | 0.045455 | 7.191288 | 5.203884 | 0.000641 | 0.027034 | 0.023911 | CHI3L1/CY | 5 |
| GO:00018 | negative regulation of cytokine production | 7/72 | 235/11391 | 0.029787 | 4.712589 | 4.586491 | 0.000685 | 0.027778 | 0.02457 | LTF/PGLYR | 7 |
| GO:19040 | positive regulation of vasculature developm | 5/72 | 112/11391 | 0.044643 | 7.062872 | 5.142522 | 0.000696 | 0.027778 | 0.02457 | CHI3L1/CY | 5 |
| GO:00485 | response to steroid hormone | 7/72 | 236/11391 | 0.029661 | 4.69262 | 4.571723 | 0.000703 | 0.027778 | 0.02457 | FOS/DEFA | 7 |
| GO:00019 | leukocyte mediated cytotoxicity | 5/72 | 115/11391 | 0.043478 | 6.878623 | 5.053254 | 0.000785 | 0.030382 | 0.026873 | TREM1/PT | 5 |
| GO:00712 | cellular response to metal ion | 5/72 | 123/11391 | 0.04065 | 6.431233 | 4.83005 | 0.001063 | 0.039525 | 0.03496 | FOS/PTGS | 5 |
| GO:00313 | positive regulation of defense response | 9/72 | 415/11391 | 0.021687 | 3.431024 | 4.023605 | 0.001154 | 0.042112 | 0.037248 | LTF/PTGS2 | 9 |

| ID | Descriptior | GeneRatio | BgRatio | RichFactor | ldEnrichme | zScore | pvalue | p.adjust | qvalue | geneID | Count |
|---|---|---|---|---|---|---|---|---|---|---|---|
| GO:00421 | T cell activ | 12/76 | 471/11391 | 0.025478 | 3.818639 | 5.120118 | 6.05E-05 | 0.045289 | 0.041727 | ZFP36L1/C | 12 |

| ID | Descriptior | GeneRatio | BgRatio | RichFactor | ldEnrichme | zScore | pvalue | p.adjust | qvalue | geneID | Count |
|---|---|---|---|---|---|---|---|---|---|---|---|

| ID | Descriptio | GeneRatic | BgRatio | RichFacto | ldEnrichm | zScore | pvalue | p.adjust | qvalue | geneID | Count |
|---|---|---|---|---|---|---|---|---|---|---|---|
| GO:00509 | leukocyte | 22/98 | 284/1139 | 0.077465 | 9.004096 | 12.72458 | 1.93E-15 | 4.13E-12 | 3.29E-12 | CXCR4/CX | 22 |
| GO:00069 | chemotax | 20/98 | 300/1139 | 0.066667 | 7.74898 | 11.03529 | 7.36E-13 | 5.24E-10 | 4.18E-10 | CXCR4/CX | 20 |
| GO:00423 | taxis | 20/98 | 300/1139 | 0.066667 | 7.74898 | 11.03529 | 7.36E-13 | 5.24E-10 | 4.18E-10 | CXCR4/CX | 20 |
| GO:00603 | cell chem | 17/98 | 209/1139 | 0.08134 | 9.454497 | 11.49136 | 1.95E-12 | 1.04E-09 | 8.3E-10 | CXCR4/CX | 17 |
| GO:00305 | leukocyte | 15/98 | 160/1139 | 0.09375 | 10.897 | 11.74423 | 5.64E-12 | 2.41E-09 | 1.92E-09 | CXCR4/CX | 15 |
| GO:00096 | response | 19/98 | 332/1139 | 0.057229 | 6.651985 | 9.736041 | 4.3E-11 | 1.45E-08 | 1.15E-08 | CXCR4/IFI | 19 |
| GO:00516 | defense re | 17/98 | 255/1139 | 0.066667 | 7.74898 | 10.15345 | 4.74E-11 | 1.45E-08 | 1.15E-08 | IFI6/IFITM | 17 |
| GO:00018 | positive re | 20/98 | 389/1139 | 0.051414 | 5.976077 | 9.302427 | 8.51E-11 | 2.27E-08 | 1.81E-08 | PTGS2/B2 | 20 |
| GO:00192 | cytokine-r | 20/98 | 395/1139 | 0.050633 | 5.885301 | 9.205402 | 1.12E-10 | 2.65E-08 | 2.12E-08 | CXCR4/CX | 20 |
| GO:00975 | myeloid le | 13/98 | 160/1139 | 0.08125 | 9.444069 | 10.02012 | 9.73E-10 | 2.08E-07 | 1.66E-07 | CXCR2/CX | 13 |
| GO:00716 | mononucl | 13/98 | 167/1139 | 0.077844 | 9.04821 | 9.760092 | 1.65E-09 | 3.21E-07 | 2.56E-07 | CXCR4/CX | 13 |
| GO:00421 | T cell activ | 20/98 | 471/1139 | 0.042463 | 4.935656 | 8.126181 | 2.45E-09 | 4.36E-07 | 3.48E-07 | B2M/ACTI | 20 |
| GO:00450 | negative r | 8/98 | 45/11391 | 0.177778 | 20.66395 | 12.31192 | 3.73E-09 | 6.12E-07 | 4.88E-07 | IFITM3/IS | 8 |
| GO:00027 | immune re | 19/98 | 448/1139 | 0.042411 | 4.929596 | 7.904765 | 6.68E-09 | 1.02E-06 | 8.12E-07 | FPR1/LAP | 19 |
| GO:00027 | immune re | 19/98 | 485/1139 | 0.039175 | 4.553524 | 7.450195 | 2.41E-08 | 3.17E-06 | 2.52E-06 | FPR1/LAP | 19 |
| GO:00022 | activation | 19/98 | 486/1139 | 0.039095 | 4.544155 | 7.438549 | 2.5E-08 | 3.17E-06 | 2.52E-06 | FPR1/LAP | 19 |
| GO:00026 | regulation | 12/98 | 172/1139 | 0.069767 | 8.109397 | 8.751655 | 2.52E-08 | 3.17E-06 | 2.52E-06 | CXCR2/DU | 12 |
| GO:00509 | regulation | 11/98 | 143/1139 | 0.076923 | 8.94113 | 8.901904 | 3.65E-08 | 4.34E-06 | 3.46E-06 | CXCR4/CX | 11 |
| GO:00507 | regulation | 10/98 | 118/1139 | 0.084746 | 9.850398 | 9.002316 | 6.31E-08 | 7.09E-06 | 5.65E-06 | CXCR4/IFI | 10 |
| GO:00485 | negative r | 8/98 | 64/11391 | 0.125 | 14.52934 | 10.11065 | 6.69E-08 | 7.15E-06 | 5.7E-06 | IFITM3/IS | 8 |
| GO:19039 | regulation | 9/98 | 95/11391 | 0.094737 | 11.01171 | 9.128047 | 1.14E-07 | 1.15E-05 | 9.21E-06 | IFITM3/IS | 9 |
| GO:00465 | host-medi | 5/98 | 15/11391 | 0.333333 | 38.7449 | 13.62634 | 1.19E-07 | 1.16E-05 | 9.23E-06 | IFITM3/LY | 5 |
| GO:00509 | positive re | 9/98 | 97/11391 | 0.092784 | 10.78466 | 9.015256 | 1.36E-07 | 1.26E-05 | 1.01E-05 | CXCR4/CX | 9 |
| GO:00450 | regulation | 8/98 | 71/11391 | 0.112676 | 13.09687 | 9.524652 | 1.53E-07 | 1.36E-05 | 1.09E-05 | IFITM3/IS | 8 |
| GO:00096 | response | 17/98 | 451/1139 | 0.037694 | 4.381352 | 6.825591 | 2.53E-07 | 2.16E-05 | 1.72E-05 | PTGS2/B2 | 17 |
| GO:00427 | defense re | 11/98 | 180/1139 | 0.061111 | 7.103231 | 7.688545 | 3.84E-07 | 3.09E-05 | 2.46E-05 | B2M/ISG1 | 11 |
| GO:00028 | regulation | 17/98 | 465/1139 | 0.036559 | 4.24944 | 6.664609 | 3.9E-07 | 3.09E-05 | 2.46E-05 | MNDA/CD | 17 |
| GO:00313 | positive re | 16/98 | 415/1139 | 0.038554 | 4.481338 | 6.730051 | 4.41E-07 | 3.37E-05 | 2.68E-05 | PTGS2/MI | 16 |
| GO:00022 | activation | 13/98 | 271/1139 | 0.04797 | 5.575834 | 7.101871 | 5.19E-07 | 3.82E-05 | 3.05E-05 | MNDA/CD | 13 |
| GO:00450 | positive re | 14/98 | 325/1139 | 0.043077 | 5.007033 | 6.827151 | 6.72E-07 | 4.78E-05 | 3.81E-05 | MNDA/CD | 14 |
| GO:00071 | leukocyte | 14/98 | 328/1139 | 0.042683 | 4.961237 | 6.781121 | 7.5E-07 | 5.17E-05 | 4.12E-05 | B2M/ACTI | 14 |
| GO:00024 | antigen pr | 5/98 | 22/11391 | 0.227273 | 26.41698 | 11.11589 | 9.97E-07 | 6.66E-05 | 5.31E-05 | B2M/MPE | 5 |
| GO:00305 | neutrophi | 7/98 | 63/11391 | 0.111111 | 12.91497 | 8.833986 | 1.05E-06 | 6.67E-05 | 5.32E-05 | CXCR2/CX | 7 |
| GO:00026 | regulation | 8/98 | 91/11391 | 0.087912 | 10.21843 | 8.224488 | 1.06E-06 | 6.67E-05 | 5.32E-05 | CXCR2/DU | 8 |
| GO:00028 | positive re | 14/98 | 342/1139 | 0.040936 | 4.758145 | 6.57348 | 1.23E-06 | 7.54E-05 | 6.01E-05 | MNDA/CD | 14 |
| GO:00450 | regulation | 15/98 | 396/1139 | 0.037879 | 4.402829 | 6.420375 | 1.32E-06 | 7.8E-05 | 6.22E-05 | MNDA/CD | 15 |
| GO:00343 | response | 8/98 | 96/11391 | 0.083333 | 9.686224 | 7.96148 | 1.6E-06 | 9.24E-05 | 7.37E-05 | IFITM3/TL | 8 |
| GO:00027 | innate imr | 12/98 | 253/1139 | 0.047431 | 5.513108 | 6.762427 | 1.66E-06 | 9.33E-05 | 7.44E-05 | CD300A/C | 12 |
| GO:00072 | positive re | 8/98 | 97/11391 | 0.082474 | 9.586367 | 7.911187 | 1.73E-06 | 9.48E-05 | 7.56E-05 | CXCR4/CX | 8 |
| GO:00421 | neutrophi | 6/98 | 44/11391 | 0.136364 | 15.85019 | 9.193649 | 1.88E-06 | 0.000101 | 8.02E-05 | CXCR2/PT | 6 |
| GO:00975 | granulocy | 8/98 | 103/1139 | 0.07767 | 9.027937 | 7.624025 | 2.73E-06 | 0.000142 | 0.000113 | CXCR2/CX | 8 |
| GO:00198 | antigen pr | 5/98 | 27/11391 | 0.185185 | 21.52494 | 9.946458 | 2.95E-06 | 0.00015 | 0.00012 | B2M/MPE | 5 |
| GO:00363 | dendritic | 5/98 | 28/11391 | 0.178571 | 20.7562 | 9.750032 | 3.57E-06 | 0.000177 | 0.000141 | CXCR4/CX | 5 |
| GO:00190 | viral geno | 8/98 | 107/1139 | 0.074766 | 8.690444 | 7.445297 | 3.64E-06 | 0.000177 | 0.000141 | IFITM3/IS | 8 |
| GO:00024 | immune re | 12/98 | 277/1139 | 0.043321 | 5.035438 | 6.333818 | 4.25E-06 | 0.000202 | 0.000161 | FPR1/LAP | 12 |
| GO:00026 | positive re | 8/98 | 111/1139 | 0.072072 | 8.377275 | 7.275673 | 4.79E-06 | 0.000223 | 0.000177 | CXCR2/PT | 8 |
| GO:00025 | monocyte | 5/98 | 30/11391 | 0.166667 | 19.37245 | 9.386196 | 5.11E-06 | 0.000229 | 0.000182 | DUSP1/FP | 5 |
| GO:00362 | granulocy | 6/98 | 52/11391 | 0.115385 | 13.4117 | 8.356331 | 5.14E-06 | 0.000229 | 0.000182 | CXCR2/PT | 6 |
| GO:00327 | positive re | 7/98 | 81/11391 | 0.08642 | 10.04497 | 7.610072 | 5.81E-06 | 0.000253 | 0.000202 | PTAFR/TL | 7 |
| GO:00190 | viral life cy | 11/98 | 239/1139 | 0.046025 | 5.349714 | 6.330726 | 6.19E-06 | 0.000259 | 0.000206 | CXCR4/IFI | 11 |
| GO:00716 | granulocy | 7/98 | 82/11391 | 0.085366 | 9.922474 | 7.553537 | 6.31E-06 | 0.000259 | 0.000206 | CXCR2/CX | 7 |
| GO:19902 | neutrophi | 7/98 | 82/11391 | 0.085366 | 9.922474 | 7.553537 | 6.31E-06 | 0.000259 | 0.000206 | CXCR2/CX | 7 |
| GO:00022 | pattern re | 11/98 | 240/1139 | 0.045833 | 5.327423 | 6.311729 | 6.44E-06 | 0.000259 | 0.000206 | CD300A/C | 11 |
| GO:19908 | response | 6/98 | 56/11391 | 0.107143 | 12.45372 | 8.003868 | 7.97E-06 | 0.000304 | 0.000242 | CXCR4/CX | 6 |
| GO:19908 | cellular re | 6/98 | 56/11391 | 0.107143 | 12.45372 | 8.003868 | 7.97E-06 | 0.000304 | 0.000242 | CXCR4/CX | 6 |
| GO:00022 | myeloid le | 10/98 | 200/1139 | 0.05 | 5.811735 | 6.395175 | 8.14E-06 | 0.000305 | 0.000243 | CXCR2/CD | 10 |
| GO:00400 | positive re | 14/98 | 406/1139 | 0.034483 | 4.008093 | 5.749426 | 9.08E-06 | 0.000334 | 0.000267 | PTGS2/CX | 14 |
| GO:00027 | immune re | 12/98 | 303/1139 | 0.039604 | 4.603354 | 5.922048 | 1.06E-05 | 0.000384 | 0.000306 | FPR1/LAP | 12 |
| GO:00327 | positive re | 6/98 | 60/11391 | 0.1 | 11.62347 | 7.685606 | 1.2E-05 | 0.000419 | 0.000334 | LILRA2/M | 6 |
| GO:00224 | regulation | 13/98 | 370/1139 | 0.035135 | 4.083922 | 5.61777 | 1.6E-05 | 0.000552 | 0.00044 | B2M/ACTI | 13 |
| GO:00480 | antigen pr | 5/98 | 39/11391 | 0.128205 | 14.90188 | 8.101026 | 1.94E-05 | 0.000648 | 0.000517 | B2M/MPE | 5 |
| GO:00026 | positive re | 6/98 | 68/11391 | 0.088235 | 10.256 | 7.131278 | 2.47E-05 | 0.0008 | 0.000638 | CXCR2/C5 | 6 |
| GO:00343 | response | 6/98 | 68/11391 | 0.088235 | 10.256 | 7.131278 | 2.47E-05 | 0.0008 | 0.000638 | IFITM3/IS | 6 |
| GO:00303 | positive re | 13/98 | 389/1139 | 0.033419 | 3.88445 | 5.392277 | 2.71E-05 | 0.000865 | 0.00069 | PTGS2/CX | 13 |
| GO:00326 | chemokin | 6/98 | 71/11391 | 0.084507 | 9.82265 | 6.946647 | 3.17E-05 | 0.000967 | 0.000771 | OAS3/TLR | 6 |
| GO:00326 | regulation | 6/98 | 71/11391 | 0.084507 | 9.82265 | 6.946647 | 3.17E-05 | 0.000967 | 0.000771 | OAS3/TLR | 6 |
| GO:20001 | positive re | 13/98 | 398/1139 | 0.032663 | 3.796611 | 5.290365 | 3.45E-05 | 0.001022 | 0.000815 | PTGS2/CX | 13 |
| GO:00507 | regulation | 11/98 | 288/1139 | 0.038194 | 4.43952 | 5.507368 | 3.54E-05 | 0.001037 | 0.000827 | PTGS2/TN | 11 |
| GO:00716 | regulation | 7/98 | 108/1139 | 0.064815 | 7.53373 | 6.355226 | 3.86E-05 | 0.001106 | 0.000882 | DUSP1/C5 | 7 |
| GO:00321 | negative r | 11/98 | 291/1139 | 0.037801 | 4.393751 | 5.463051 | 3.9E-05 | 0.001106 | 0.000882 | TNFRSF1B | 11 |
| GO:00069 | phagocyto | 9/98 | 192/1139 | 0.046875 | 5.448501 | 5.790885 | 3.93E-05 | 0.001106 | 0.000882 | CD93/CD3 | 9 |
| GO:19030 | regulation | 11/98 | 296/1139 | 0.037162 | 4.319533 | 5.390504 | 4.55E-05 | 0.001263 | 0.001007 | B2M/ACTI | 11 |
| GO:00071 | G protein- | 14/98 | 476/1139 | 0.029412 | 3.418667 | 5.021552 | 5.31E-05 | 0.00144 | 0.001148 | CXCR4/CX | 14 |
| GO:00712 | cellular re | 9/98 | 200/1139 | 0.045 | 5.230561 | 5.62275 | 5.41E-05 | 0.00144 | 0.001148 | PTGS2/B2 | 9 |
| GO:00198 | antigen pr | 6/98 | 78/11391 | 0.076923 | 8.94113 | 6.555578 | 5.42E-05 | 0.00144 | 0.001148 | B2M/MPE | 6 |
| GO:00507 | positive re | 7/98 | 114/1139 | 0.061404 | 7.137218 | 6.134758 | 5.46E-05 | 0.00144 | 0.001148 | PTGS2/TL | 7 |
| GO:00015 | blood ves | 14/98 | 479/1139 | 0.029228 | 3.397256 | 4.993445 | 5.69E-05 | 0.001482 | 0.001181 | PTGS2/CX | 14 |
| GO:00508 | regulation | 11/98 | 306/1139 | 0.035948 | 4.178371 | 5.250102 | 6.15E-05 | 0.001583 | 0.001262 | B2M/ACTI | 11 |
| GO:01403 | antiviral ir | 5/98 | 51/11391 | 0.098039 | 11.39556 | 6.931011 | 7.3E-05 | 0.001857 | 0.001481 | OAS3/OAS | 5 |
| GO:00075 | blood coa | 8/98 | 162/1139 | 0.049383 | 5.739985 | 5.660231 | 7.49E-05 | 0.001881 | 0.0015 | SERPINA1 | 8 |
| GO:00508 | coagulatic | 8/98 | 165/1139 | 0.048485 | 5.635622 | 5.587373 | 8.52E-05 | 0.002089 | 0.001666 | SERPINA1 | 8 |
| GO:00712 | cellular re | 8/98 | 165/1139 | 0.048485 | 5.635622 | 5.587373 | 8.52E-05 | 0.002089 | 0.001666 | PTGS2/B2 | 8 |
| GO:00326 | interleukir | 5/98 | 53/11391 | 0.09434 | 10.96554 | 6.773929 | 8.8E-05 | 0.002089 | 0.001666 | LAPTM5/T | 5 |
| GO:00326 | regulation | 5/98 | 53/11391 | 0.09434 | 10.96554 | 6.773929 | 8.8E-05 | 0.002089 | 0.001666 | LAPTM5/T | 5 |
| GO:00327 | positive re | 5/98 | 53/11391 | 0.09434 | 10.96554 | 6.773929 | 8.8E-05 | 0.002089 | 0.001666 | LILRA2/M | 5 |
| GO:00075 | hemostasi | 8/98 | 167/1139 | 0.047904 | 5.568129 | 5.539786 | 9.27E-05 | 0.002139 | 0.001706 | SERPINA1 | 8 |
| GO:00326 | interleukir | 7/98 | 124/1139 | 0.056452 | 6.561636 | 5.800688 | 9.31E-05 | 0.002139 | 0.001706 | PTAFR/TL | 7 |
| GO:00326 | regulation | 7/98 | 124/1139 | 0.056452 | 6.561636 | 5.800688 | 9.31E-05 | 0.002139 | 0.001706 | PTAFR/TL | 7 |
| GO:00327 | positive re | 5/98 | 54/11391 | 0.092593 | 10.76247 | 6.698504 | 9.63E-05 | 0.002179 | 0.001738 | OAS3/TLR | 5 |
| GO:00302 | T cell diffe | 10/98 | 272/1139 | 0.036765 | 4.273334 | 5.089934 | 0.000113 | 0.002503 | 0.001996 | B2M/ACTI | 10 |
| GO:00712 | cellular re | 8/98 | 174/1139 | 0.045977 | 5.344124 | 5.379089 | 0.000123 | 0.002717 | 0.002166 | PTGS2/B2 | 8 |
| GO:00713 | cellular re | 13/98 | 452/1139 | 0.028761 | 3.343033 | 4.735099 | 0.000126 | 0.002745 | 0.002189 | PTGS2/B2 | 13 |
| GO:00326 | interleukir | 6/98 | 92/11391 | 0.065217 | 7.580524 | 5.903432 | 0.000137 | 0.002887 | 0.002302 | LILRA2/M | 6 |
| GO:00326 | regulation | 6/98 | 92/11391 | 0.065217 | 7.580524 | 5.903432 | 0.000137 | 0.002887 | 0.002302 | LILRA2/M | 6 |
| GO:00160 | viral proce | 11/98 | 341/1139 | 0.032258 | 3.749506 | 4.801987 | 0.000161 | 0.003371 | 0.002688 | CXCR4/IFI | 11 |
| GO:00026 | regulation | 13/98 | 466/1139 | 0.027897 | 3.242599 | 4.604729 | 0.00017 | 0.003501 | 0.002792 | B2M/ACTI | 13 |
| GO:00508 | defense re | 5/98 | 61/11391 | 0.081967 | 9.527434 | 6.22069 | 0.000173 | 0.003516 | 0.002803 | B2M/TLR2 | 5 |
| GO:00603 | type I inte | 5/98 | 63/11391 | 0.079365 | 9.224976 | 6.098156 | 0.000201 | 0.004021 | 0.003207 | IFITM3/IS | 5 |
| GO:00485 | blood ves | 12/98 | 413/1139 | 0.029056 | 3.377279 | 4.584203 | 0.000213 | 0.004128 | 0.003292 | PTGS2/CX | 12 |
| GO:00512 | regulation | 12/98 | 413/1139 | 0.029056 | 3.377279 | 4.584203 | 0.000213 | 0.004128 | 0.003292 | B2M/ACTI | 12 |
| GO:00713 | cellular re | 5/98 | 64/11391 | 0.078125 | 9.080835 | 6.038917 | 0.000217 | 0.004177 | 0.00333 | IFITM3/IS | 5 |
| GO:01500 | neuroinfla | 5/98 | 66/11391 | 0.075758 | 8.805659 | 5.92424 | 0.000251 | 0.004744 | 0.003783 | PTGS2/TN | 5 |
| GO:00224 | positive re | 9/98 | 249/1139 | 0.036145 | 4.201254 | 4.757825 | 0.000285 | 0.005255 | 0.00419 | B2M/ACTI | 9 |
| GO:19016 | response | 7/98 | 149/1139 | 0.04698 | 5.46069 | 5.105563 | 0.000291 | 0.005319 | 0.004241 | TXNIP/PTA | 7 |
| GO:00015 | angiogene | 11/98 | 367/1139 | 0.029973 | 3.483874 | 4.505709 | 0.000304 | 0.005504 | 0.004388 | PTGS2/CX | 11 |
| GO:00519 | regulation | 7/98 | 155/1139 | 0.045161 | 5.249309 | 4.961906 | 0.00037 | 0.006594 | 0.005258 | PTGS2/CX | 7 |
| GO:00072 | phospholi | 5/98 | 72/11391 | 0.069444 | 8.071854 | 5.607454 | 0.000377 | 0.00666 | 0.005311 | CXCR2/FP | 5 |
| GO:19031 | mononucl | 12/98 | 443/1139 | 0.027088 | 3.14857 | 4.296889 | 0.000403 | 0.007059 | 0.005628 | B2M/ACTI | 12 |
| GO:00313 | negative r | 8/98 | 209/1139 | 0.038278 | 4.449175 | 4.688121 | 0.00043 | 0.007444 | 0.005935 | TNFRSF1B | 8 |
| GO:00622 | regulation | 7/98 | 159/1139 | 0.044025 | 5.117251 | 4.870209 | 0.000432 | 0.007444 | 0.005935 | CD300A/C | 7 |
| GO:00432 | regulation | 9/98 | 267/1139 | 0.033708 | 3.918023 | 4.494529 | 0.000476 | 0.008066 | 0.006431 | PTGS2/B2 | 9 |
| GO:00022 | response | 9/98 | 268/1139 | 0.033582 | 3.903404 | 4.480579 | 0.000489 | 0.008222 | 0.006556 | PTGS2/B2 | 9 |
| GO:00027 | cytoplasm | 7/98 | 165/1139 | 0.042424 | 4.931169 | 4.738287 | 0.00054 | 0.009007 | 0.007182 | OAS3/OAS | 7 |
| GO:00971 | intrinsic a | 9/98 | 274/1139 | 0.032847 | 3.817928 | 4.398267 | 0.000574 | 0.009499 | 0.007574 | PTGS2/TN | 9 |
| GO:00326 | interleukir | 5/98 | 81/11391 | 0.061728 | 7.174981 | 5.195377 | 0.00065 | 0.0106 | 0.008452 | LILRA2/M | 5 |
| GO:00326 | regulation | 5/98 | 81/11391 | 0.061728 | 7.174981 | 5.195377 | 0.00065 | 0.0106 | 0.008452 | LILRA2/M | 5 |
| GO:00026 | negative r | 11/98 | 406/1139 | 0.027094 | 3.149216 | 4.107838 | 0.000711 | 0.01143 | 0.009114 | LAPTM5/D | 11 |
| GO:00071 | adenylate | 6/98 | 125/1139 | 0.048 | 5.579265 | 4.795525 | 0.000715 | 0.01143 | 0.009114 | FPR1/FPR | 6 |
| GO:19037 | regulation | 10/98 | 343/1139 | 0.029155 | 3.388767 | 4.184554 | 0.000717 | 0.01143 | 0.009114 | B2M/ACTI | 10 |
| GO:01408 | interferon | 5/98 | 84/11391 | 0.059524 | 6.918732 | 5.071832 | 0.000768 | 0.012055 | 0.009612 | IFITM3/IS | 5 |
| GO:00512 | negative r | 6/98 | 129/1139 | 0.046512 | 5.406265 | 4.688435 | 0.000844 | 0.013071 | 0.010422 | LAPTM5/M | 6 |
| GO:00518 | host-medi | 5/98 | 87/11391 | 0.057471 | 6.680155 | 4.954205 | 0.0009 | 0.013834 | 0.011031 | IFITM3/LY | 5 |
| GO:00109 | regulation | 8/98 | 237/1139 | 0.033755 | 3.923534 | 4.236793 | 0.000981 | 0.01476 | 0.011769 | PTGS2/B2 | 8 |
| GO:00420 | wound he | 9/98 | 300/1139 | 0.03 | 3.487041 | 4.066572 | 0.001093 | 0.016147 | 0.012875 | CXCR4/SE | 9 |
| GO:00620 | regulation | 8/98 | 242/1139 | 0.033058 | 3.842469 | 4.163473 | 0.001122 | 0.016398 | 0.013075 | PTGS2/PT | 8 |
| GO:00300 | lymphocy | 10/98 | 364/1139 | 0.027473 | 3.193261 | 3.961709 | 0.001129 | 0.016398 | 0.013075 | B2M/ACTI | 10 |
| GO:00508 | regulation | 8/98 | 248/1139 | 0.032258 | 3.749506 | 4.078023 | 0.001313 | 0.018822 | 0.015008 | SERPINA1 | 8 |
| GO:00358 | modulatio | 5/98 | 96/11391 | 0.052083 | 6.05389 | 4.632213 | 0.001401 | 0.019957 | 0.015913 | IFITM3/LY | 5 |
| GO:00324 | response | 8/98 | 252/1139 | 0.031746 | 3.68999 | 4.022519 | 0.001454 | 0.020563 | 0.016396 | PTGS2/B2 | 8 |
| GO:00023 | leukocyte | 8/98 | 253/1139 | 0.031621 | 3.675405 | 4.008819 | 0.001491 | 0.020947 | 0.016703 | PTAFR/CD | 8 |
| GO:00022 | cell activa | 8/98 | 256/1139 | 0.03125 | 3.632334 | 3.968132 | 0.001606 | 0.022133 | 0.017648 | PTAFR/CD | 8 |
| GO:00301 | platelet ac | 5/98 | 100/1139 | 0.05 | 5.811735 | 4.502002 | 0.00168 | 0.022572 | 0.017998 | ACTB/IL6F | 5 |
| GO:00508 | negative r | 5/98 | 100/1139 | 0.05 | 5.811735 | 4.502002 | 0.00168 | 0.022572 | 0.017998 | LAPTM5/C | 5 |
| GO:00305 | intracellul | 9/98 | 323/1139 | 0.027864 | 3.238738 | 3.802245 | 0.001822 | 0.024026 | 0.019157 | OAS3/OAS | 9 |
| GO:00096 | response | 10/98 | 390/1139 | 0.025641 | 2.980377 | 3.7071 | 0.001888 | 0.024636 | 0.019644 | CXCR4/SE | 10 |
| GO:00107 | positive re | 9/98 | 325/1139 | 0.027692 | 3.218807 | 3.780386 | 0.001901 | 0.024636 | 0.019644 | CXCR4/AC | 9 |
| GO:00022 | T cell activ | 5/98 | 103/1139 | 0.048544 | 5.642461 | 4.408883 | 0.001915 | 0.024636 | 0.019644 | ITGAL/FGI | 5 |
| GO:00316 | receptor i | 5/98 | 103/1139 | 0.048544 | 5.642461 | 4.408883 | 0.001915 | 0.024636 | 0.019644 | CXCR2/CX | 5 |
| GO:00607 | regulation | 6/98 | 152/1139 | 0.039474 | 4.588212 | 4.148645 | 0.001963 | 0.025113 | 0.020024 | CXCR4/LA | 6 |
| GO:00027 | regulation | 5/98 | 104/1139 | 0.048077 | 5.588206 | 4.378654 | 0.001998 | 0.025401 | 0.020254 | B2M/TNFI | 5 |
| GO:00026 | negative r | 6/98 | 153/1139 | 0.039216 | 4.558223 | 4.127667 | 0.00203 | 0.025652 | 0.020454 | LAPTM5/M | 6 |
| GO:00023 | cytokine p | 5/98 | 105/1139 | 0.047619 | 5.534985 | 4.348813 | 0.002084 | 0.02618 | 0.020875 | B2M/TNFI | 5 |
| GO:00068 | receptor-r | 7/98 | 209/1139 | 0.033493 | 3.893028 | 3.932206 | 0.002137 | 0.026388 | 0.021041 | B2M/CXCI | 7 |
| GO:19030 | negative r | 5/98 | 106/1139 | 0.04717 | 5.482769 | 4.319353 | 0.002172 | 0.026664 | 0.021261 | LAPTM5/C | 5 |
| GO:00072 | Rho prote | 5/98 | 109/1139 | 0.045872 | 5.331867 | 4.233168 | 0.002454 | 0.029448 | 0.02348 | TAGAP/AI | 5 |
| GO:00024 | leukocyte | 9/98 | 338/1139 | 0.026627 | 3.095007 | 3.642285 | 0.002479 | 0.029587 | 0.023591 | B2M/TNFI | 9 |
| GO:00620 | positive re | 5/98 | 113/1139 | 0.044248 | 5.143128 | 4.12308 | 0.00287 | 0.033364 | 0.026603 | PTGS2/PT | 5 |
| GO:19021 | positive re | 6/98 | 164/1139 | 0.036585 | 4.252489 | 3.908194 | 0.002874 | 0.033364 | 0.026603 | ACTB/EGF | 6 |
| GO:19037 | positive re | 6/98 | 164/1139 | 0.036585 | 4.252489 | 3.908194 | 0.002874 | 0.033364 | 0.026603 | ACTB/EGF | 6 |
| GO:00016 | response | 7/98 | 221/1139 | 0.031674 | 3.681642 | 3.75008 | 0.002923 | 0.033749 | 0.02691 | PTGS2/CX | 7 |
| GO:19030 | positive re | 7/98 | 223/1139 | 0.03139 | 3.648623 | 3.72096 | 0.003073 | 0.034366 | 0.027402 | B2M/ACTI | 7 |
| GO:00456 | regulation | 6/98 | 168/1139 | 0.035714 | 4.151239 | 3.833114 | 0.003239 | 0.035475 | 0.028286 | B2M/ISG1 | 6 |
| GO:00508 | negative r | 6/98 | 169/1139 | 0.035503 | 4.126676 | 3.814708 | 0.003335 | 0.036343 | 0.028978 | LAPTM5/M | 6 |
| GO:00082 | negative r | 11/98 | 494/1139 | 0.022267 | 2.588222 | 3.361951 | 0.003381 | 0.036654 | 0.029227 | PTGS2/B2 | 11 |
| GO:00362 | response | 7/98 | 230/1139 | 0.030435 | 3.537578 | 3.621612 | 0.003645 | 0.039323 | 0.031355 | PTGS2/CX | 7 |
| GO:00457 | positive re | 9/98 | 359/1139 | 0.02507 | 2.913962 | 3.432603 | 0.003703 | 0.039571 | 0.031552 | B2M/ACTI | 9 |
| GO:00971 | apoptotic | 11/98 | 500/1139 | 0.022 | 2.557163 | 3.317077 | 0.003705 | 0.039571 | 0.031552 | PTGS2/TN | 11 |
| GO:00301 | regulation | 7/98 | 231/1139 | 0.030303 | 3.522263 | 3.607735 | 0.003733 | 0.039671 | 0.031632 | B2M/ACTI | 7 |
| GO:00466 | lymphocy | 7/98 | 232/1139 | 0.030172 | 3.507081 | 3.593933 | 0.003823 | 0.039723 | 0.031673 | TNFRSF1B | 7 |
| GO:00300 | myeloid c | 9/98 | 364/1139 | 0.024725 | 2.873935 | 3.384907 | 0.004054 | 0.041636 | 0.033199 | B2M/ISG1 | 9 |
| GO:00485 | response | 7/98 | 236/1139 | 0.029661 | 3.447639 | 3.539475 | 0.004197 | 0.04249 | 0.03388 | PTGS2/TX | 7 |
| GO:00434 | regulation | 10/98 | 439/1139 | 0.022779 | 2.647715 | 3.279729 | 0.004419 | 0.044525 | 0.035503 | LAPTM5/D | 10 |
| GO:00080 | blood circ | 8/98 | 302/1139 | 0.02649 | 3.079065 | 3.411108 | 0.004441 | 0.044537 | 0.035512 | PTGS2/CX | 8 |
| GO:00329 | mononucl | 7/98 | 239/1139 | 0.029289 | 3.404363 | 3.499395 | 0.004496 | 0.044587 | 0.035552 | TNFRSF1B | 7 |
| GO:00508 | positive re | 8/98 | 304/1139 | 0.026316 | 3.058808 | 3.389345 | 0.00462 | 0.044796 | 0.035718 | B2M/ACTI | 8 |
| GO:00430 | positive re | 10/98 | 442/1139 | 0.022624 | 2.629744 | 3.255469 | 0.004635 | 0.044796 | 0.035718 | PTGS2/TX | 10 |
| GO:00507 | negative r | 5/98 | 127/1139 | 0.03937 | 4.576169 | 3.775233 | 0.004732 | 0.045321 | 0.036137 | TNFRSF1B | 5 |
| GO:00026 | regulation | 8/98 | 312/1139 | 0.025641 | 2.980377 | 3.304038 | 0.005391 | 0.049482 | 0.039455 | B2M/TNFI | 8 |
